# Comparative efficacy and safety of targeted disease-modifying therapies in adults with primary IgA nephropathy: a systematic review and network meta-analysis

**DOI:** 10.64898/2026.10.25.26366409

**Authors:** Zijian Liu, Yue Wang, Mingzhu Zhang, Manuel Sanchez, Yuchen Fu, Huiting Sun, Luying Sun

**Affiliations:** Department of Nephrology and Endocrinology, Dongzhimen Hospital, Beijing University of Chinese Medicine, Beijing 100700, China; Department of Nephrology, Fangshan Hospital, Beijing University of Chinese Medicine, Beijing 102400, China; Faculty of Medicine, National Autonomous University of Mexico (UNAM), Mexico City, Mexico

**Keywords:** IgA nephropathy, targeted therapies, network meta-analysis, proteinuria, estimated glomerular filtration rate

## Abstract

**Background:** Targeted disease-modifying therapies for adults with primary IgA nephropathy are increasing in number. However, head-to-head evidence is limited, and trial endpoints and follow-up durations vary. The relative effects of these therapies on short-term proteinuria control, long-term kidney outcomes, and safety remain unclear.

**Methods:** We searched databases and trial registries for randomized, double-blind, placebo-controlled trials with a planned treatment duration of ≥24 weeks alongside stable or optimized supportive care. Frequentist random-effects network meta-analyses evaluated baseline-adjusted geometric mean ratios (GMRs) for 24-hour urinary protein-to-creatinine ratio at 32–40 and 24–52 weeks, with Holm correction applied. Total estimated glomerular filtration rate (eGFR) slope and composite kidney outcomes were synthesized using common-effect models. Serious adverse events (SAEs) were analyzed using penalized-likelihood logistic regression within risk windows. Sensitivity analyses included restriction to evidence at low risk of bias, exclusion of interim or stage-specific reports, and assessment of precision-recovery assumptions and time-window boundaries. The Mantel–Haenszel method without continuity correction assessed the robustness of sparse SAE results. RoB 2 and CINeMA assessed risk of bias and evidence certainty, respectively.

**Results:** Seventeen trials were included. At 32–40 weeks, telitacicept and subcutaneous sibeprenlimab had the lowest GMR point estimates versus placebo: 0.450 (nominal 95% CI 0.323–0.628) and 0.488 (0.355–0.671), respectively. None of the seven targeted treatment comparisons reached statistical significance after Holm correction. Iptacopan had the largest point estimate for improvement in annualized total eGFR slope: 3.020 mL/min/1.73 m^2^/year (95% CI 2.025–4.015). Composite kidney outcome hazard ratios were 0.570 (95% CI 0.401– 0.811) for iptacopan and 0.440 (0.271–0.714) for targeted-release budesonide. Telitacicept had the lowest SAE odds ratio point estimate at 36–39 weeks: 0.314 (95% CI 0.105–0.938), although safety estimates were imprecise. In sensitivity analyses, proteinuria effect directions were generally consistent, but the 95% CIs for all retained comparisons included 1 after excluding interim or stage-specific reports; sparse SAE estimates remained imprecise. Evidence certainty was rated as low or very low.

**Conclusion:** Although some regimens yielded the most favorable point estimates, the available evidence remains insufficient to establish treatment priorities or comparative safety advantages. Head-to-head trials are needed.

**Systematic review registration:** https://www.crd.york.ac.uk/PROSPERO/,identifier CRD420261358501.

## 1. Introduction

Primary IgA nephropathy can cause progressive loss of kidney function and kidney failure (1). Supportive care, including renin–angiotensin system (RAS) blockade and sodium–glucose cotransporter 2 (SGLT2) inhibitors, is the foundation of disease management. Some patients nevertheless have persistent proteinuria and remain at risk of progression despite optimized treatment (2, 3). Gut-targeted-release budesonide, selective endothelin A receptor antagonists, complement inhibitors, and APRIL and/or BAFF pathway inhibitors act at different stages of pathogenesis. Several regimens have reduced proteinuria, and some have also provided evidence on long-term kidney function or composite kidney outcomes (3–6).

Existing trials primarily compare targeted therapies with placebo, and head-to-head evidence remains lacking (7). Differences in study populations, background supportive care, and follow-up time points, together with uneven evidence coverage of short-term proteinuria, long-term kidney outcomes, and safety, complicate clinical treatment selection. We conducted a systematic review and network meta-analysis of aggregate data from randomized controlled trials to compare proteinuria, kidney function, composite kidney outcomes, and SAEs across specific drugs and dosing regimens alongside stable or optimized supportive care, and to assess evidence certainty at the comparison level.

## 2 Methods

### 2.1 Study design and reporting standards

This systematic review and frequentist network meta-analysis used aggregate data from randomized controlled trials to compare the efficacy and safety of targeted therapies alongside stable or optimized supportive care. The review was registered in PROSPERO (CRD420261358501) and is reported in accordance with the PRISMA 2020 statement and the PRISMA-NMA extension (8, 9).

### 2.2 Population and eligibility criteria

We included randomized, double-blind, parallel-group, placebo-controlled trials in adults aged ≥18 years with biopsy-confirmed primary IgA nephropathy who received stable or optimized RAS inhibition. The planned randomized treatment period had to be at least 24 weeks. We excluded patients with secondary IgA nephropathy, IgA vasculitis–associated kidney disease, a predominantly crescentic or rapidly progressive glomerulonephritis phenotype, or a kidney transplant. Crossover trials, single-arm trials, nonrandomized and observational studies, and active-comparator trials without a placebo arm were excluded.

### 2.3 Interventions and comparators

Interventions were targeted drugs acting on pathogenic pathways in IgA nephropathy: gut-targeted-release budesonide; selective endothelin A receptor antagonists (atrasentan and SC0062); complement pathway inhibitors (ravulizumab, cemdisiran, iptacopan, narsoplimab, and vemircopan); APRIL and/or BAFF pathway inhibitors (telitacicept, sibeprenlimab, and atacicept); and the anti-CD38 monoclonal antibody felzartamab. Treatment nodes were defined by active ingredient, formulation, route of administration, and dose–frequency regimen. Comparators were the concurrent matched placebo arms of each trial, with both groups receiving the background supportive care specified in the original protocol. We extracted requirements for RAS inhibitor optimization and actual use, SGLT2 inhibitor use, restrictions on immunosuppression, and rescue treatment. Placebo arms across trials formed the common reference node.

### 2.4 Search and study selection

We systematically searched PubMed, Embase, Web of Science Core Collection, the Cochrane Central Register of Controlled Trials (CENTRAL), ClinicalTrials.gov, and the WHO International Clinical Trials Registry Platform (ICTRP) on October 15, 2026, from inception to the search date. Supplementary searches covered systematic reviews, conference abstracts, and reference lists of included articles. Two reviewers independently screened titles and abstracts and assessed full texts. Disagreements were resolved through discussion or, when necessary, adjudication by a third reviewer. We extracted study design, treatment regimens, background therapies, baseline characteristics, and outcome data. Eligibility criteria, outcome selection rules, and complete search strategies are provided in Supplementary Material Appendices 1–3.

### 2.5 Statistical analysis

We performed frequentist network meta-analyses, synthesizing proteinuria, kidney function, and composite kidney outcomes using a contrast-based, inverse-variance-weighted graph-theoretical approach (10). One target treatment arm and its concurrent placebo control from each independently randomized cohort were included for each outcome and time window. Proteinuria was expressed as a baseline-adjusted GMR, annualized total eGFR slope as a mean difference (MD), and composite kidney outcomes as an HR. GMRs and HRs were synthesized on the logarithmic scale and back-transformed. All effects were reported with two-sided 95% confidence intervals (CIs).

Separate proteinuria networks were constructed for the 32–40-week primary window and the 24– 52-week broad window. Random-effects models assumed a common between-study variance (τ^2^) across comparisons, estimated by restricted maximum likelihood (REML). CIs used a t distribution, with degrees of freedom equal to the number of independent study contrasts minus the rank of the network design matrix. Holm correction was applied to multiple testing of primary-window active treatment–placebo comparisons (11). The 95% CIs were nominal intervals without multiplicity adjustment, and the two-sided significance level was 0.05. The kidney function and composite kidney outcome networks lacked replicated direct comparisons, making between-study variance unidentifiable; common-effect models with normal-based 95% CIs were therefore used. Comparability of composite kidney outcomes was assessed by event components and sustained-confirmation rules, and outcomes representing similar clinical constructs underwent exploratory synthesis.

For SAEs, inputs were the number of participants experiencing at least one event and the corresponding safety analysis denominator. Separate networks were constructed for approximate 24–26-week and 36–39-week risk windows according to the actual event collection periods. A common-effect Firth penalized-likelihood logistic regression model with study-specific intercepts was fitted (12), and ORs with normal-based Wald 95% CIs were reported. Sensitivity analyses used the Mantel–Haenszel method without continuity correction (13).

Sensitivity analyses restricted studies to those at low risk of bias for the relevant outcome, excluded interim or stage-specific reports and results at time-window boundaries, and sequentially omitted individual studies for direct comparisons supported by multiple studies. Additional scenario analyses fixed heterogeneity parameters and varied assumptions about degrees of freedom in the original trials. None of the outcome networks contained a closed loop that allowed incoherence testing. Transitivity was therefore assessed qualitatively by comparing potential effect modifiers, including baseline proteinuria, eGFR, background treatment, and assessment time points (14). Results were interpreted using effect size, precision, and CINeMA evidence certainty; treatments were not ranked. Analyses were conducted in R 4.4.1 using netmeta 3.6.1 and meta 8.5.0 (15).

### 2.6 Risk of bias and evidence certainty

Two reviewers independently used the Cochrane RoB 2 tool (16) to assess individual trial outcome results across five domains: the randomization process, deviations from intended interventions, missing outcome data, outcome measurement, and selection of the reported result. Overall risk of bias was classified as low risk, some concerns, or high risk; disagreements were resolved through discussion. Trial registrations, protocols, and public reports informed the ROB-MEN assessment of bias due to missing evidence in network comparisons (17). The CINeMA framework (18) assessed the certainty of treatment comparisons for each outcome across six domains: within-study bias, reporting bias, indirectness, imprecision, heterogeneity, and incoherence. Certainty was rated as high, moderate, low, or very low. Shared concerns across related domains were considered jointly when downgrading to avoid double counting, and results were interpreted in relation to effect size and precision.

## 3 Results

### 3.1 Search and study selection

Following screening and full-text assessment, 17 randomized, double-blind, placebo-controlled trials were included: two evaluated targeted-release budesonide (3, 19), two selective endothelin A receptor antagonists (4, 20), six complement pathway inhibitors (5, 21–25), six APRIL and/or BAFF pathway inhibitors (6, 26–30), and one an anti-CD38 monoclonal antibody (31). Each trial compared targeted therapy with concurrent placebo alongside protocol-specified supportive care. The study selection process is shown in Figure 1.

**Figure 1.**
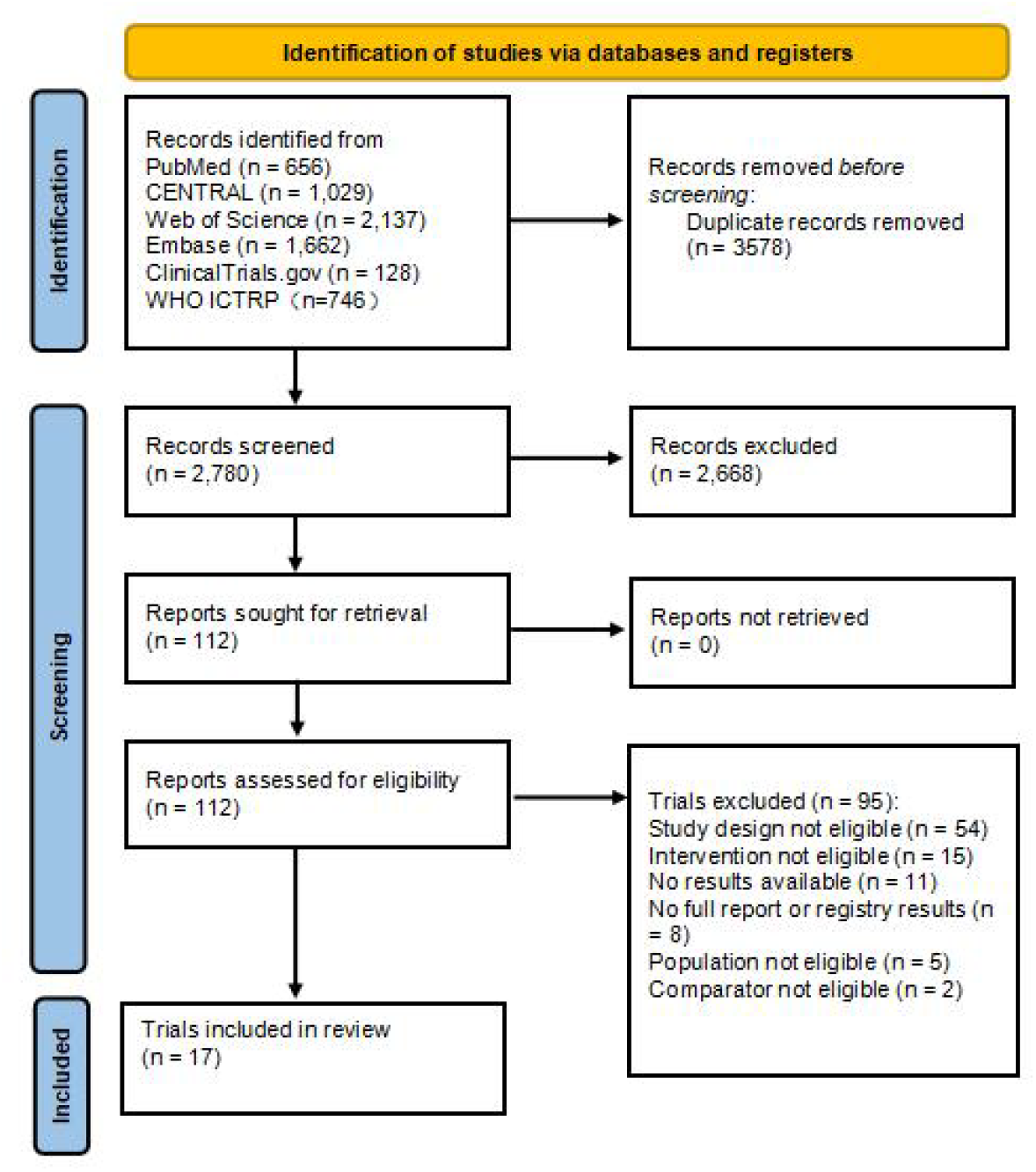
Literature search and study selection flow diagram.

### 3.2 Study and patient characteristics

The included trials investigated 12 targeted drugs across Europe, North America, the Asia-Pacific region, and South America. Most were multinational, while some were conducted in China.

Among target treatment arms and concurrent controls with reported baseline data, mean or median age ranged from 36.0 to 45.8 years, and women accounted for 23.1%–66.7%. All trials were conducted alongside stable or optimized RAS blockade. SGLT2 inhibitor use ranged from 0% to 55.7% in arms with reported data. Baseline characteristics and background therapies are presented in Supplementary Table 4.1.

### 3.3 Risk of bias

RoB 2 assessments covered 25 individual outcome results: four were at low risk, 18 had some concerns, and three were at high risk. Of 13 proteinuria results, four were at low risk and nine had some concerns. Of three annualized total eGFR slope results, two were at high risk and one had some concerns. Both composite kidney outcome results had some concerns. Of five SAE results, one was at high risk and four had some concerns. High-risk judgments for long-term eGFR slope primarily reflected censoring of observations associated with disease progression and potentially missing information in some trials. For SAEs, the principal concern was exclusion of randomized participants who received no study drug, whose influence on sparse-event effect estimates could not be ruled out. In the missing outcome data domain, 19 results had some concerns and two were at high risk. All 25 results were at low risk in the outcome measurement domain. After ROB-MEN reassessment, no missing evidence related to the results was identified for direct treatment– placebo comparisons, which were judged at low risk; active treatment comparisons retained some concerns. Result-specific assessments and assessments of bias due to missing evidence are provided in Supplementary Tables 6.1–6.3.

### 3.4 Efficacy outcomes

The primary proteinuria window (32–40 weeks) included nine trials and 2,218 participants. GMRs versus placebo were 0.599 (95% CI 0.412–0.872) for atacicept, 0.617 (0.468–0.814) for atrasentan, 0.626 (0.182–2.156) for cemdisiran, and 0.610 (0.466–0.798) for iptacopan.

Corresponding GMRs for subcutaneous sibeprenlimab, targeted-release budesonide, and telitacicept were 0.488 (0.355–0.671), 0.704 (0.542–0.914), and 0.450 (0.323–0.628), respectively. Nominal 95% CIs excluded 1 for all regimens except cemdisiran; none of the seven treatment–placebo comparisons reached statistical significance after Holm correction.

The broad proteinuria window (24–52 weeks) included 13 trials and 2,451 participants. GMRs versus placebo were 0.450 (95% CI 0.352–0.576) for telitacicept and 0.488 (0.386–0.618) for subcutaneous sibeprenlimab. Estimates for the additional regimens, ravulizumab, SC0062, and intravenous sibeprenlimab, were 0.668 (0.450–0.991), 0.484 (0.297–0.789), and 0.516 (0.271– 0.980), respectively. Among indirect active treatment comparisons, GMRs for telitacicept versus iptacopan and targeted-release budesonide were 0.728 (0.532–0.996) and 0.639 (0.468–0.874), respectively; the GMR for subcutaneous sibeprenlimab versus targeted-release budesonide was 0.693 (0.511–0.940). The 95% CIs for all other comparisons included 1.

The annualized total eGFR slope analysis included three trials and 1,181 participants, with assessment at 104 or 136 weeks. MDs versus placebo were 1.410 (95% CI 0.489–2.331) for atrasentan, 3.020 (2.025–4.015) for iptacopan, and 1.820 (0.505–3.135) mL/min/1.73 m^2^/year for targeted-release budesonide. The MD for iptacopan versus atrasentan was 1.610 (0.254–2.966) mL/min/1.73 m^2^/year; intervals for the remaining active treatment comparisons included 0. The exploratory composite kidney outcome analysis included two trials and 841 participants, both with 104 weeks of follow-up. HRs versus placebo were 0.570 (95% CI 0.401–0.811) for iptacopan and 0.440 (0.271–0.714) for targeted-release budesonide; the HR for their indirect comparison was 1.295 (0.712–2.357). Relative effects and evidence certainty for the main outcomes are presented in Table 1. Direct evidence is provided in Supplementary Material Appendix 5, complete pairwise comparisons in Supplementary Tables 7.2–7.7, and network and forest plots in Supplementary Figures 7.1–7.6.

**Table 1.** Treatment effects versus placebo and CINeMA certainty of evidence across outcome-specific analysis windows.

| Treatment regimen | Proteinuria<br>32–40 weeks<br>GMR (95% CI) | Proteinuria<br>24–52 weeks<br>GMR (95% CI) | Total eGFR<br>slope<br>MD (95% CI) | Kidney<br>composite<br>104 weeks<br>HR (95% CI) | SAEs<br>24–26 weeks<br>OR (95% CI) | SAEs<br>36–39 weeks<br>OR (95% CI) |
| --- | --- | --- | --- | --- | --- | --- |
| Atacicept 150 mg<br>SC weekly | 0.599**<br>(0.412–0.872) | 0.599**<br>(0.454–0.791) | — | — | — | 0.415*<br>(0.056–3.081) |
| Atrasentan 0.75 mg<br>oral daily | 0.617*<br>(0.468–0.814) | 0.617*<br>(0.503–0.757) | 1.410*<br>(0.489–2.331) | — | — | — |
| Cemdisiran 600 mg<br>SC every 4 weeks | 0.626*<br>(0.182–2.156) | 0.626*<br>(0.251–1.562) | — | — | — | 1.326*<br>(0.042–41.440) |
| Iptacopan 200 mg<br>oral twice daily | 0.610*<br>(0.466–0.798) | 0.618*<br>(0.509–0.750) | 3.020*<br>(2.025–4.015) | 0.570*<br>(0.401–0.811) | — | — |
| Ravulizumab<br>weight-based IV | — | 0.668**<br>(0.450–0.991) | — | — | 1.659*<br>(0.061–45.047) | — |
| SC0062 20 mg<br>oral daily | — | 0.484*<br>(0.297–0.789) | — | — | 1.893*<br>(0.228–15.696) | — |
| Sibeprenlimab 400 mg<br>SC every 4 weeks | 0.488**<br>(0.355–0.671) | 0.488**<br>(0.386–0.618) | — | — | — | — |
| Sibeprenlimab 4 mg/kg<br>IV every 4 weeks | — | 0.516*<br>(0.271–0.980) | — | — | — | — |
| Targeted-release<br>budesonide 16 mg<br>oral daily | 0.704*<br>(0.542–0.914) | 0.704*<br>(0.580–0.854) | 1.820*<br>(0.505–3.135) | 0.440*<br>(0.271–0.714) | — | — |
| Telitacicept 240 mg<br>SC weekly | 0.450**<br>(0.323–0.628) | 0.450**<br>(0.352–0.576) | — | — | — | 0.314*<br>(0.105–0.938) |

All estimates compare the listed regimen with placebo; all groups received trial-specified background supportive care. GMR, HR, or OR <1 and eGFR slope MD >0 favor the listed regimen. A dash indicates no estimate in that analysis set.
CINeMA certainty: \*\*low; \*very low. Asterisks indicate evidence certainty, not statistical significance. Proteinuria CIs are nominal; all seven primary- window treatment–placebo comparisons had Holm-adjusted $P > 0.05$ . Kidney function and composite outcomes used exploratory common-effect models; SAEs used Firth penalized-likelihood logistic models. Complete pairwise comparisons are reported in Supplementary Tables 7.2–7.7.
GMR, geometric mean ratio; MD, mean difference; HR, hazard ratio; OR, odds ratio; CI, confidence interval; eGFR, estimated glomerular filtration rate; SAE, serious adverse event; SC, subcutaneous; IV, intravenous. Total eGFR slope MDs are expressed in mL/min/1.73 m<sup>2</sup>/year.

### 3.5 Serious adverse events

The 24–26-week and 36–39-week risk windows included two and three trials, respectively. The 95% CIs included 1 for most treatment–placebo comparisons and all active treatment comparisons. The OR for telitacicept versus placebo at 36–39 weeks was 0.314 (95% CI 0.105– 0.938). However, sparse events, limited observation, and very low evidence certainty precluded confirmation of a safety advantage. Raw event counts, collection periods, and comparisons between analytical methods are provided in Supplementary Material Appendix 9 and Supplementary Table 8.5.

### 3.6 Sensitivity analyses

Restriction to studies with proteinuria results at low RoB 2 risk retained three trials and 841 participants in the primary window and four trials and 907 participants in the broad window, covering three and four active regimens, respectively. All retained regimens had point estimates below 1 versus control.

After excluding the interim reports by Perkovic 2026 (VISIONARY) and Lafayette 2026 (ORIGIN phase 3), seven trials remained in the primary window. The subcutaneous sibeprenlimab node was removed, and the atacicept GMR changed from 0.599 to 0.650. Point estimates for other retained nodes were largely unchanged, and all treatment–control intervals crossed 1. Eleven trials remained in the broad window; intervals for atacicept, ravulizumab, and intravenous sibeprenlimab changed from excluding to crossing 1. Further exclusion of the prespecified 39-week stage-specific TELIGAN analysis made the telitacicept node inestimable because no other eligible evidence remained. Point estimates for the other retained regimens changed little. After excluding results at the 32- and 40-week boundaries, the cemdisiran and subcutaneous sibeprenlimab nodes were removed; point estimates and intervals for the remaining nodes were largely unchanged.

For direct comparisons supported by at least two trials, sequential omission of one trial and reanalysis yielded GMRs of 0.700–0.717 for budesonide and 0.582–0.650 for atacicept. The corresponding 95% CIs in the primary window all included 1. In the broad window, excluding Barratt 2026 (APPLAUSE-IgAN) changed the iptacopan GMR from 0.618 to 0.800 (95% CI 0.239–2.678); excluding Zhang 2024 (the phase 2 iptacopan trial) yielded a GMR of 0.610 (0.466–0.798).

Proteinuria point estimates remained below 1 after changes to between-study heterogeneity assumptions and CI calculation methods. Sensitivity and additional scenario analyses are provided in Supplementary Material Appendix 8.

### 3.7 Evidence certainty

Of 28 comparisons in the primary proteinuria window, three had low and 25 had very low certainty; the corresponding numbers among 55 broad-window comparisons were four and 51. Low-certainty comparisons were atacicept, subcutaneous sibeprenlimab, and telitacicept versus placebo, with ravulizumab versus placebo additionally included in the broad window. All active treatment comparisons and all comparisons for the remaining outcomes had very low certainty.

The low-certainty comparisons were primarily limited by the absence of closed loops, which prevented incoherence testing. Limited estimability of heterogeneity was considered jointly with this structural limitation when downgrading. Other direct proteinuria comparisons retained concerns about within-study bias, and cemdisiran had major concerns about imprecision in both windows. Active treatment comparisons were additionally affected by cross-trial differences in baseline risk, background SGLT2 inhibitor use, and handling of intercurrent events. Major imprecision concerns applied to 14 of 21 indirect comparisons in the primary window and 31 of 45 in the broad window.

For annualized total eGFR slope, comparisons involving atrasentan and targeted-release budesonide had major concerns about within-study bias because of censoring associated with disease progression. The two direct composite kidney outcome comparisons were downgraded primarily for within-study bias and the network structural limitations described above. Their indirect comparison was additionally affected by differences in endpoint confirmation rules and intercurrent event strategies, and had major concerns about imprecision. All SAE comparisons except telitacicept versus placebo at 36–39 weeks had major concerns about imprecision.

Comparison-level judgments across the six domains and overall ratings are provided in Supplementary Tables 6.4–6.9.

## 4 Discussion

### 4.1 Strengths and limitations

This study has several strengths. It addresses targeted treatment selection alongside stable or optimized supportive care, separately synthesizing short-term proteinuria, long-term eGFR trajectories, and composite kidney outcomes to distinguish early treatment responses from long-term kidney benefits. For sparse SAEs, Firth penalized-likelihood logistic regression mitigated separation caused by zero events and small-sample estimation bias; the Mantel–Haenszel method without continuity correction assessed sensitivity to the analytical method. Given the sparse evidence networks and predominance of indirect comparisons, treatments were not ranked using SUCRA or P-scores. Instead, results were interpreted using effect size, precision, and evidence certainty. RoB 2, ROB-MEN, and CINeMA assessed within-study bias, bias due to missing evidence, and comparison-level evidence certainty, respectively. Sensitivity analyses addressing interim or stage-specific reports, assumptions used to recover precision, and time-window boundaries examined the dependence of the main inferences on specific studies and analytical assumptions.

The principal limitations are sparse evidence networks and limited cross-trial comparability. There were no head-to-head trials of active treatments and no closed loops permitting incoherence testing; the validity of indirect comparisons therefore depends on the transitivity assumption. Trials differed in baseline kidney function, proteinuria, background SGLT2 inhibitor use, and rescue treatment management. Residual differences may affect relative effect estimates despite harmonization of outcomes and assessment windows. Long-term eGFR slope and composite kidney outcomes were supported by few trials, without independent replication of the corresponding direct comparisons. Composite endpoints also differed in event components, sustained-confirmation rules, and handling of intercurrent events. SAE synthesis was limited by event counts and observation duration, reducing the ability to identify rare, delayed, and drug-specific risks. Reliance on reported aggregate data also limited assessment of patient-level differences in treatment effects. The findings primarily apply to adults with primary IgA nephropathy whose characteristics resemble those of the included trial populations. Extrapolation to advanced kidney disease, low-proteinuria populations, and combination or sequential treatment requires corresponding evidence.

### 4.2 Comparison with previous evidence

Previous network meta-analyses have produced different proteinuria rankings under different eligibility criteria and effect scales. Chen et al. synthesized 24-hour urinary protein excretion using standardized mean differences and reported the highest SUCRA for telitacicept (32); telitacicept also had the lowest GMR point estimate in our primary window. Mansour et al. focused on the BAFF/APRIL pathway and reported the highest proteinuria P-score for sibeprenlimab, although between-drug differences were not statistically significant (33). These analyses addressed different questions: the former included conventional immunosuppression, supportive therapies, and novel targeted drugs, whereas the latter compared individual molecules within a specified pathway. Our study restricted eligibility to targeted therapies alongside stable or optimized supportive care, distinguished specific dosing regimens, synthesized proteinuria using GMRs, and separately assessed short-term proteinuria, annualized total eGFR slope over at least 104 weeks, composite kidney outcomes, and safety.

For safety, the CIs for all active treatment SAE comparisons in our study crossed 1, consistent with previous findings that sparse evidence cannot support reliable safety rankings (7).

Telitacicept had the lowest SAE OR point estimate versus placebo in the 36–39-week risk window, but events were few and the estimate remained uncertain. Differences in observation periods, dose pooling, and safety analysis populations may influence these sparse-event estimates. Clinical interpretation should consider both overall SAEs and drug-specific risks.

### 4.3 Mechanisms of action and clinical implications

In this study, telitacicept and subcutaneous sibeprenlimab had relatively favorable proteinuria effect point estimates. Iptacopan had the largest point estimate for improvement in annualized total eGFR slope, and both iptacopan and targeted-release budesonide showed signals of benefit for composite kidney outcomes. These treatments act on pathogenic IgA production and the intrarenal inflammatory responses it triggers. Deposition of IgA immune complexes can activate mesangial cells and promote podocyte injury, glomerulosclerosis, and tubulointerstitial fibrosis through the release of proinflammatory and profibrotic mediators (34). In vitro experiments using patient-derived IgA1 complexes further showed that CXCL1 and TGF-β1 secreted by activated mesangial cells synergistically increased podocyte death and impaired adhesion (35). The biological effects of targeted therapies thus involve ongoing immune injury and its cumulative consequences, while long-term changes in kidney function are also influenced by pre-existing chronic lesions (34).

Both telitacicept and sibeprenlimab intervene in APRIL-related IgA production. APRIL promotes IgA class switching in mucosal B cells through TACI and supports plasma cell maintenance through TACI- and BCMA-related signaling; BAFF also regulates B-cell survival and differentiation (36). Telitacicept neutralizes both BAFF and APRIL, whereas sibeprenlimab selectively inhibits APRIL. Both act on upstream immune processes involved in IgA production (36). An ancillary phase 2 telitacicept study observed reductions in galactose-deficient IgA1 (Gd-IgA1) and IgA-containing immune complexes that correlated with proteinuria improvement (37). The phase 3 sibeprenlimab trial also reported reduced Gd-IgA1 levels (28).

In this study, iptacopan had the largest point estimate for improvement in annualized total eGFR slope and showed a signal of benefit for composite kidney outcomes. Iptacopan targets complement factor B, inhibiting alternative pathway C3 convertase activity and complement amplification. This reduces downstream C5a and C5b-9 generation and limits inflammation and complement-mediated cellular injury (23, 34). Injury involving complement activation extends beyond the filtration barrier to mesangial inflammation, podocyte injury, and tubulointerstitial inflammation (34). These processes collectively promote glomerulosclerosis, interstitial fibrosis, and nephron loss and are closely associated with progressive loss of kidney function (34). The phase 2 trial observed reductions in plasma Bb and urinary soluble C5b-9, together with suppression of functional alternative pathway activity (23). APPLAUSE-IgAN further showed that suppression of functional alternative pathway activity and reductions in urinary soluble C5b-9 were sustained through 24 months (5).

Targeted-release budesonide showed signals of benefit for annualized total eGFR slope and composite kidney outcomes in this study. Its distal ileal delivery is designed to modulate IgA immune responses in gut-associated lymphoid tissue (3, 34). The NEFIGAN biomarker analysis observed reductions in circulating Gd-IgA1 and IgA–IgG immune complexes (34, 38).

Deposition of these complexes can activate mesangial cells and induce proinflammatory and profibrotic responses that contribute to progressive kidney injury (34). In NefIgArd, the between-group eGFR difference established during 9 months of treatment was maintained during the subsequent 15-month off-treatment follow-up, and a reduced risk of composite kidney outcomes was observed (3).

Clinical decisions should integrate persistent proteinuria, longitudinal eGFR changes, previous treatment responses, and long-term outcome evidence for each regimen. Local approved indications, drug availability, treatment costs, patient preferences, and drug-specific risks must also be considered. For example, the administration burden and long-term adherence requirements of once-weekly subcutaneous telitacicept and twice-daily oral iptacopan should inform shared decision-making. Telitacicept requires attention to changes in immunoglobulin levels and associated infection risk; iptacopan requires vaccination against encapsulated bacteria, antimicrobial prophylaxis when indicated, and infection monitoring in accordance with local prescribing requirements (6, 39). Follow-up should assess both proteinuria response and the kidney function trajectory to evaluate the short-term response and progress toward long-term kidney protection.

## 5 Conclusion

### Conclusion

In the 32–40-week primary proteinuria window, telitacicept and subcutaneous sibeprenlimab had the largest point estimates for proteinuria reduction. Synthesis of long-term outcomes indicated that iptacopan had the largest point estimate for improvement in annualized total eGFR slope; both iptacopan and targeted-release budesonide showed signals of benefit for composite kidney outcomes. Telitacicept had the lowest SAE OR point estimate in the 36–39-week risk window. However, the certainty of the available evidence is insufficient to establish a clear treatment hierarchy, and SAE estimates were generally imprecise. High-quality head-to-head randomized controlled trials are needed to compare the relative efficacy and safety of these agents.

## Data Availability

The data that support the findings of this study are available from the corresponding author upon reasonable request.

## Data availability statement

The original contributions presented in this study are included in the article and Supplementary Material. Further inquiries can be directed to the corresponding author.

## Author contributions

ZL: Conceptualization, Data curation, Formal analysis, Investigation, Methodology, Software, Writing – original draft, Writing – review & editing. YW: Conceptualization, Formal analysis, Investigation, Writing – original draft, Writing – review & editing. MZ: Validation, Visualization, Writing – original draft, Writing – review & editing. MS: Formal analysis, Project administration, Supervision, Validation, Visualization, Writing – original draft, Writing – review & editing. YF: Investigation, Methodology, Project administration, Writing – original draft, Writing – review & editing. HS: Data curation, Methodology, Writing – original draft, Writing – review & editing.LS: Resources, Supervision, Validation, Writing – original draft, Writing – review & editing.

## Generative AI statement

The author(s) declared that generative AI was not used in the creation of this manuscript.

---

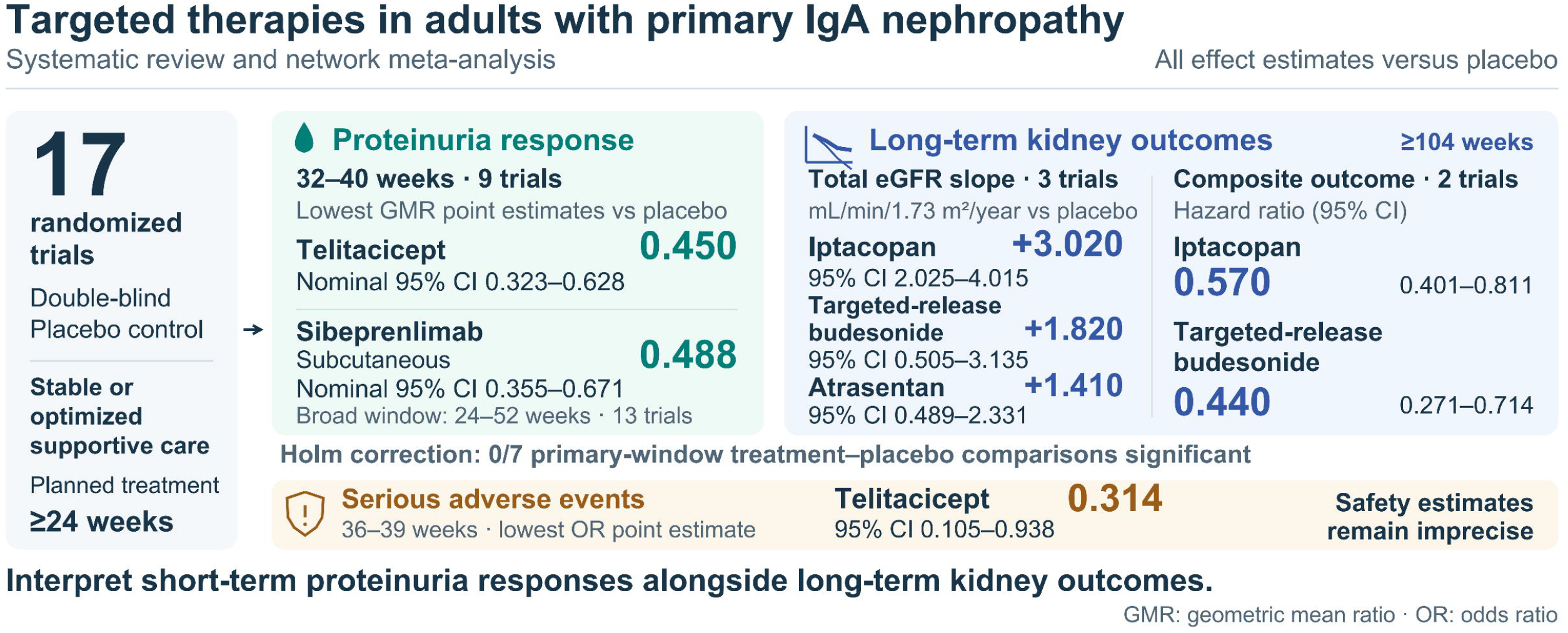

