## Supplementary_Material for "Comparative efficacy and safety of targeted disease-modifying therapies in adults with primary IgA nephropathy: a systematic review and network meta-analysis"

**Supplementary Material for the Network Meta-analysis of Targeted Therapies for IgA Nephropathy**

**Contents**

APPENDIX 1 Study Scope and Analysis Plan	2

APPENDIX 2 Detailed Methods and Analysis Sets	3

APPENDIX 3 Search Strategies	6

APPENDIX 4 Study Characteristics and Transitivity	7

APPENDIX 5 Direct Evidence and Pairwise Analyses	10

APPENDIX 6 Risk of Bias and Certainty of Evidence	14

APPENDIX 7 Detailed Network Meta-analysis Results	22

APPENDIX 8 Sensitivity and Supplementary Analyses	32

APPENDIX 9 Raw Serious Adverse Event Data and Risk Windows	35

APPENDIX 10 Subsequent Trials and Long-term Follow-up	39

Methodological References	42

### **APPENDIX 1 Study Scope and Analysis Plan**

#### **1.1 Research Question and Eligibility Criteria**

To evaluate the efficacy of different targeted therapies relative to concurrent placebo and explore relative effects between active treatments in the setting of stable or optimized renin–angiotensin system (RAS) blockade.

**Table 1.1 Research question and scope of inclusion**

| **Domain** | **Element** | **Operational definition** |
| --- | --- | --- |
| Study population | Population | Adults aged ≥18 years with biopsy-confirmed primary IgA nephropathy. |
| Study comparisons | Intervention and comparator | Targeted therapy versus matching placebo; each group received background supportive care specified in the original trial. |
| Study design | Design and treatment duration | Randomized, double-blind, parallel-group controlled trials with a planned randomized treatment period of at least 24 weeks. |
| Study eligibility | Exclusions | Nonrandomized or crossover trials; secondary IgA nephropathy, IgA vasculitis, transplant recipients, or rapidly progressive phenotypes; and mixed populations for which IgA nephropathy results could not be extracted separately. |

#### **1.2 Treatment Nodes and Study Units**

Treatment nodes were defined by active ingredient, formulation, route of administration, dose, and dosing frequency.

#### **1.3 Outcome Hierarchy and Result Selection**

The primary outcome was the baseline-adjusted geometric mean ratio (GMR) for the urine protein-to-creatinine ratio (UPCR) measured in a complete 24-hour urine collection at 32–40 weeks (inclusive). Measurement type, analysis population, estimand, and precision eligibility were checked before selecting the result closest to week 36; the later time point was selected when two time points were equally distant.

Proteinuria at 24–52 weeks was examined in a time-window sensitivity analysis. Annualized total eGFR slope, composite kidney outcomes, and serious adverse events (SAEs) were exploratory outcomes.

Total eGFR slope was required to start at randomization, include the acute phase, and cover at least 104 weeks of observation.

Composite outcomes were distinguished by their components, confirmation rules, and intercurrent event strategies. SAEs were synthesized separately within the 24–26-week and 36–39-week risk windows.

#### **1.4 Analysis Plan and Presentation of Results**

The primary proteinuria analysis focused on the effects of active treatments relative to placebo. Supplementary analyses examined the effects of the time window, precision recovery, risk of bias, interim reports, and direct comparisons supported by multiple studies. Results were presented as network plots, effect estimates with 95% confidence intervals, direct comparisons, and certainty-of-evidence assessments. SUCRA or P-score rankings were not reported because the evidence was sparse and comparisons between active treatments relied predominantly on indirect evidence.

### **APPENDIX 2 Detailed Methods and Analysis Sets**

#### **2.1 Literature Search and Study Identification**

PubMed, Embase, Web of Science Core Collection, the Cochrane Central Register of Controlled Trials (CENTRAL), ClinicalTrials.gov, and the WHO International Clinical Trials Registry Platform (ICTRP) were searched from inception to the search date. Supplementary searches covered systematic reviews, conference abstracts, and the reference lists of included articles. Search strategies combined controlled vocabulary with free-text terms; the full strategies are provided in APPENDIX 3.

#### **2.2 Data Extraction and Background Treatment**

Data were extracted on study design, study regions, eligibility criteria, numbers randomized and analyzed, treatment regimens, follow-up, age, sex, race, baseline eGFR, proteinuria, blood pressure, disease duration, and prior immunosuppression. For background treatment, RAS optimization requirements, actual medication use, SGLT2 inhibitor use, and restrictions on immunosuppression were recorded separately.

For continuous outcomes, adjusted between-group estimates, confidence intervals or standard errors, measurement time points, and the original statistical models were extracted; for binary outcomes, the numbers of participants with events and the corresponding analysis denominators were extracted.

#### **2.3 Effect Measures and Precision**

GMRs were used for proteinuria, hazard ratios (HRs) for composite kidney outcomes, and odds ratios (ORs) for SAEs; all were analyzed on the logarithmic scale. Mean differences (MDs) were used for eGFR outcomes; total slope was expressed in mL/min/1.73 m²/year. UPCR reported in g/mol or mg/mmol was converted to g/g by dividing by 113.12.

#### **2.4 Transitivity and Risk of Bias**

Potential effect modifiers were examined by direct comparison, including baseline eGFR and proteinuria, age, blood pressure, region and race, background RAS and SGLT2 inhibitor use, restrictions on immunosuppression, treatment exposure, and intercurrent event strategies. Characteristics of the target analysis populations were used to assess comparability. RoB 2 was applied to specific results across five domains: randomization, deviations from intended interventions, missing outcome data, outcome measurement, and selection of the reported result.[3]

#### **2.5 Statistical Synthesis and Multiplicity**

A frequentist inverse-variance network meta-analysis was performed. The proteinuria model assumed a common heterogeneity variance across comparisons, estimated by restricted maximum likelihood (REML); intervals were based on the t distribution, with degrees of freedom equal to the number of independent contrasts minus the rank of the design matrix. A common-effect model was used as a reference. For direct comparisons supported by at least two studies, inverse-variance common-effect pairwise synthesis with normal-based intervals was performed; I² was calculated from Cochran’s Q, and τ² was estimated separately by REML. Pairwise heterogeneity was not estimated for single-study comparisons. The current kidney function and composite outcome networks had no residual degrees of freedom; common-effect models were therefore used to provide conditional estimates.

Statistical analyses were performed using R 4.4.1, netmeta 3.6.1, and meta 8.5.0.

Active treatment–placebo comparisons for the primary proteinuria outcome constituted one family of comparisons, with Holm adjustment applied to P values from the corresponding t tests; the 95% confidence intervals were nominal. Comparisons between active treatments, as well as analyses of other outcomes, were interpreted as exploratory. Inconsistency between direct and indirect evidence could not be tested because the networks contained no closed loops.

#### **2.6 Composite Kidney Outcomes and Safety**

The original trial HRs, event components, and requirements for sustained confirmation were retained for composite kidney outcomes, and exploratory synthesis was undertaken for sufficiently similar constructs. For SAEs, the numerator was the number of participants experiencing at least one event and the denominator was the original safety risk set; risk windows were defined according to the actual collection periods. A common-effect penalized-likelihood logistic regression (LRP) model was used, with the Mantel–Haenszel (MH) method without continuity correction used for comparison.

#### **2.7 Sensitivity Analyses and Certainty of Evidence**

Sensitivity analyses included the 24–52-week broad proteinuria window, restriction to results at low risk of bias according to RoB 2, and exclusion of interim reports or reports at the time-window boundaries. For direct comparisons supported by at least two trials, individual trials were excluded in turn and the network model was refitted. Robustness was assessed by varying fixed heterogeneity values and the degrees of freedom assumed for the original trials. CINeMA was used to assess within-study bias, reporting bias, indirectness, imprecision, heterogeneity, and incoherence.[5] For direct treatment–placebo comparisons, cross-checking trial registrations and available reports identified no evidence missing on the basis of the results; ROB-MEN judgments were low risk, and no downgrading was applied in the CINeMA reporting bias domain.[4,7] Incoherence was consistently judged to raise major concerns when there were no closed loops and a global test could not be performed. Limited estimability of heterogeneity and this structural limitation were considered jointly when downgrading, without counting them twice.[5]

#### **2.8 Study Results and Analysis Sets**

**Table 2.1 Study results and analysis sets**

| **Study** | **Outcome** | **Assessment time point or risk window** | **Analysis set** |
| --- | --- | --- | --- |
| Fellström 2017（NEFIGAN） | Proteinuria (24-hour UPCR) | 39 weeks | Primary window; broad window |
| Lafayette 2023（NefIgArd） | Proteinuria (24-hour UPCR) | 39 weeks | Primary window; broad window |
| Lafayette 2023（NefIgArd） | Annualized total eGFR slope | 104 weeks | Exploratory total eGFR slope analysis |
| Lafayette 2023（NefIgArd） | Composite kidney outcome | 104 weeks | Exploratory composite outcome analysis |
| Heerspink 2026（ALIGN） | Proteinuria (24-hour UPCR) | 36 weeks | Primary window; broad window |
| Heerspink 2026（ALIGN） | Annualized total eGFR slope | 136 weeks | Exploratory total eGFR slope analysis |
| Heerspink 2025（SC0062） | Proteinuria (24-hour UPCR) | 24 weeks | Broad window |
| Heerspink 2025（SC0062） | Participants with at least one SAE | 24 weeks | Exploratory early-window SAE analysis |
| Lafayette 2025（SANCTUARY） | Proteinuria (24-hour UPCR) | 26 weeks | Broad window |
| Lafayette 2025（SANCTUARY） | Participants with at least one SAE | 26 weeks | Exploratory early-window SAE analysis |
| Barratt 2024（Cemdisiran） | Proteinuria (24-hour UPCR) | 32 weeks | Primary window; broad window |
| Barratt 2024（Cemdisiran） | Participants with at least one SAE | 36 weeks | Exploratory late-window SAE analysis |
| Barratt 2026（APPLAUSE-IgAN） | Proteinuria (24-hour UPCR) | 39 weeks | Primary window; broad window |
| Barratt 2026（APPLAUSE-IgAN） | Annualized total eGFR slope | 104 weeks | Exploratory total eGFR slope analysis |
| Barratt 2026（APPLAUSE-IgAN） | Composite kidney outcome | 104 weeks | Exploratory composite outcome analysis |
| Mathur 2024（ENVISION） | Proteinuria (24-hour UPCR) | 52 weeks | Broad window |
| Perkovic 2026（VISIONARY） | Proteinuria (24-hour UPCR) | 40 weeks | Primary window; broad window |
| Lafayette 2024 (ORIGIN phase IIb) | Proteinuria (24-hour UPCR) | 36 weeks | Primary window; broad window |
| Lafayette 2024 (ORIGIN phase IIb) | Participants with at least one SAE | 36 weeks | Exploratory late-window SAE analysis |
| Lafayette 2026 (ORIGIN phase III) | Proteinuria (24-hour UPCR) | 36 weeks | Primary window; broad window |
| Lv 2026（TELIGAN） | Proteinuria (24-hour UPCR) | 39 weeks | Primary window; broad window |
| Lv 2026（TELIGAN） | Participants with at least one SAE | 39 weeks | Exploratory late-window SAE analysis |
| Zhang 2024 (Iptacopan phase II) | Proteinuria (24-hour UPCR) | 25.7 weeks | Broad window |

Note: Primary proteinuria window: 32–40 weeks. Broad proteinuria window: 24-52 weeks

### **APPENDIX 3 Search Strategies**

#### **3.1 PubMed**

(("Glomerulonephritis, IGA"[Mesh] OR ((IgA[tiab] OR "immunoglobulin A"[tiab]) AND (nephropath*[tiab] OR glomerulonephrit*[tiab] OR nephrit*[tiab])) OR IgAN[tiab] OR "Berger disease"[tiab] OR "Berger's disease"[tiab]) AND ("randomized controlled trial"[pt] OR "controlled clinical trial"[pt] OR randomized[tiab] OR placebo[tiab] OR drug therapy[sh] OR randomly[tiab] OR trial[tiab] OR groups[tiab]) NOT (animals[mh] NOT humans[mh]))

#### **3.2 CENTRAL**

(MeSH descriptor: [Glomerulonephritis, IGA] explode all trees) OR (((IgA NEAR/3 (nephropath* OR glomerulonephrit* OR nephrit*)) OR ("immunoglobulin A" NEAR/3 (nephropath* OR glomerulonephrit* OR nephrit*)) OR IgAN OR "Berger disease" OR "Berger's disease"):ti,ab,kw)

#### **3.3 Web of Science Core Collection**

TS=((IgA NEAR/3 (nephropath* OR glomerulonephrit* OR nephrit*)) OR ("immunoglobulin A" NEAR/3 (nephropath* OR glomerulonephrit* OR nephrit*)) OR IgAN OR "Berger disease" OR "Berger's disease") AND TS=(random* OR placebo* OR trial* OR ((single OR double OR triple OR treble) NEAR/2 (blind* OR mask*)) OR crossover OR "cross over" OR (controlled NEAR/3 (study OR studies)))

#### **3.4 Embase**

('immunoglobulin a glomerulonephritis'/exp OR (((iga NEAR/3 (nephropath* OR glomerulonephrit* OR nephrit*)) OR ('immunoglobulin a' NEAR/3 (nephropath* OR glomerulonephrit* OR nephrit*)) OR igan OR 'berger disease' OR "berger's disease"):ti,ab,kw,tt)) AND (('randomized controlled trial'/exp OR 'controlled clinical trial'/de OR random*:ti,ab,tt OR 'randomization'/de OR 'intermethod comparison'/de OR placebo:ti,ab,tt OR (compare:ti,tt OR compared:ti,tt OR comparison:ti,tt) OR ((evaluated:ab OR evaluate:ab OR evaluating:ab OR assessed:ab OR assess:ab) AND (compare:ab OR compared:ab OR comparing:ab OR comparison:ab)) OR (open NEXT/1 label):ti,ab,tt OR ((double OR single OR doubly OR singly) NEXT/1 (blind OR blinded OR blindly)):ti,ab,tt OR 'double blind procedure'/de OR (parallel NEXT/1 group*):ti,ab,tt OR (crossover:ti,ab,tt OR 'cross over':ti,ab,tt) OR ((assign* OR match OR matched OR allocation) NEAR/6 (alternate OR group OR groups OR intervention OR interventions OR patient OR patients OR subject OR subjects OR participant OR participants)):ti,ab,tt OR (assigned:ti,ab,tt OR allocated:ti,ab,tt) OR (controlled NEAR/8 (study OR design OR trial)):ti,ab,tt OR (volunteer:ti,ab,tt OR volunteers:ti,ab,tt) OR 'human experiment'/de OR trial:ti,tt) NOT ((((random* NEXT/1 sampl* NEAR/8 ('cross section*' OR questionnaire* OR survey OR surveys OR database OR databases)):ti,ab,tt) NOT ('comparative study'/de OR 'controlled study'/de OR 'randomised controlled':ti,ab,tt OR 'randomized controlled':ti,ab,tt OR 'randomly assigned':ti,ab,tt)) OR ('cross-sectional study'/de NOT ('randomized controlled trial'/exp OR 'controlled clinical study'/de OR 'controlled study'/de OR 'randomised controlled':ti,ab,tt OR 'randomized controlled':ti,ab,tt OR 'control group':ti,ab,tt OR 'control groups':ti,ab,tt)) OR ('case control*':ti,ab,tt AND random*:ti,ab,tt NOT ('randomised controlled':ti,ab,tt OR 'randomized controlled':ti,ab,tt)) OR ('systematic review':ti,tt NOT (trial:ti,tt OR study:ti,tt)) OR (nonrandom*:ti,ab,tt NOT random*:ti,ab,tt) OR 'random field*':ti,ab,tt OR ('random cluster' NEAR/4 sampl*):ti,ab,tt OR ((review:ab AND review:it) NOT trial:ti,tt) OR ('we searched':ab AND (review:ti,tt OR review:it)) OR 'update review':ab OR (databases NEAR/5 searched):ab OR ((rat:ti,tt OR rats:ti,tt OR mouse:ti,tt OR mice:ti,tt OR swine:ti,tt OR porcine:ti,tt OR murine:ti,tt OR sheep:ti,tt OR lambs:ti,tt OR pigs:ti,tt OR piglets:ti,tt OR rabbit:ti,tt OR rabbits:ti,tt OR cat:ti,tt OR cats:ti,tt OR dog:ti,tt OR dogs:ti,tt OR cattle:ti,tt OR bovine:ti,tt OR monkey:ti,tt OR monkeys:ti,tt OR trout:ti,tt OR marmoset*:ti,tt) AND 'animal experiment'/de) OR ('animal experiment'/de NOT ('human experiment'/de OR 'human'/de))))

#### **3.5 ClinicalTrials.gov**

"IgA nephropathy" OR "IgA nephritis" OR "IgA glomerulonephritis" OR "immunoglobulin A nephropathy" OR "immunoglobulin A nephritis" OR "immunoglobulin A glomerulonephritis" OR "Berger disease" OR "Berger's disease" OR IgAN

#### **3.6 WHO ICTRP**

"IgA nephropathy" OR "IgA nephritis" OR "IgA glomerulonephritis" OR "immunoglobulin A nephropathy" OR "immunoglobulin A nephritis" OR "immunoglobulin A glomerulonephritis" OR "Berger disease" OR "Berger's disease" OR IgAN

### **APPENDIX 4 Study Characteristics and Transitivity**

#### **4.1 Target Treatment Arms and Concurrent Controls**

**Table 4.1 Key baseline characteristics of target treatment arms and concurrent controls**

| **Study** | **Study-arm treatment and dose** | **Age (years)** | **Female ( %)** | **Baseline eGFR  (mL/min/1.73 m²)** | **Baseline proteinuria,  UPCR (g/g)¹** | **Baseline background treatment,  RASi/SGLT2i (%)** | **Immunosuppressant use** | **Participa nts per arm (n)** |
| --- | --- | --- | --- | --- | --- | --- | --- | --- |
| **a Primary proteinuria window, 32–40 weeks (9 RCTs)** | | | | | | | | |
| Fellström 2017 NEFIGAN | Targeted-release budesonide 16 mg  orally, once daily | 37.5 (11.9) | 31.2 | 83.8 (25.9) | 0.8 [0.5–1.3] | RASi 100.0 ; SGLT2i not obtained | Systemic immunosuppression for >1 week during the preceding 24 months was an exclusion criterion; prohibited during the trial. | 48 |
|  | Concurrent matching placebo | 38.9 (12) | 30.0 | 76.5 (23.2) | 0.8 [0.5–1.6] | RASi 100.0 ; SGLT2i not obtained | Systemic immunosuppression for >1 week during the preceding 24 months was an exclusion criterion; prohibited during the trial. | 50 |
| Lafayette 2023 NefIgArd | Targeted-release budesonide 16 mg  orally, once daily | 43 [36–50] | 35.7 | 56.14 [45.5–70.97] | 1.28 [0.9–1.76] | RASi 98.4 ; SGLT2i not obtained | Immunosuppression restricted before randomization | 182 |
|  | Concurrent matching placebo | 42 [34–49] | 32.4 | 55.11 [45.96–67.74] | 1.25 [0.88–1.74] | RASi 98.4 ; SGLT2i not obtained | Immunosuppression restricted before randomization | 182 |
| Heerspink 2026 ALIGN | Atrasentan 0.75 mg  orally, once daily | 45.8 (12.7) | 39.4 | 58.1 (23.6) | 1.4 [1–2] | RASi 99.4 SGLT2i 0.0 | Prohibited medications and rescue treatment managed according to protocol; pre-enrollment washout period unclear. | 170 |
|  | Concurrent matching placebo | 43.7 (11.2) | 45.3 | 59.7 (24.3) | 1.4 [1.1–1.9] | RASi 98.2 SGLT2i 0.0 | Prohibited medications and rescue treatment managed according to protocol; pre-enrollment washout period unclear. | 170 |
| Barratt 2024 Cemdisiran | Cemdisiran 600 mg  subcutaneously, every 4 weeks | 41 (10) | 40.9 | 68 [54–94] | 1.55 (1.03) | ACEi 31.8；ARB 63.6c SGLT2i 0.0 | Immunosuppressive treatment within the preceding 6 months was an exclusion criterion. | 22 |
|  | Concurrent matching placebo | 38 (10) | 66.7 | 47 [39–76] | 1.97 (0.82) | ACEi 11.1；ARB 88.9c SGLT2i 0.0 | Immunosuppressive treatment within the preceding 6 months was an exclusion criterion. | 9 |
| Barratt 2026 APPLAUSE-IgAN | Iptacopan 200 mg  orally, twice daily | 39.7 (11.6) | 43.7 | 63.4 (25.5) | 1.7 [1.3–2.4] | RASi 100.0 SGLT2i 21.4 | Immunosuppressive or immunomodulatory treatment within 90 days before the first dose was an exclusion criterion. | 238 |
|  | Concurrent matching placebo | 39.2 (12.5) | 49.4 | 65.6 (26.5) | 1.8 [1.5–2.6] | RASi 98.3 SGLT2i 17.2 | Immunosuppressive or immunomodulatory treatment within 90 days before the first dose was an exclusion criterion. | 239 |
| Perkovic 2026 VISIONARY | Sibeprenlimab 400 mg  subcutaneously, every 4 weeks | 42 [18–75]a | 34.2 | 63.49 (24.4) | 1.22 [0.49–6.69]a | RASi 98.0 SGLT2i 36.8 | Current systemic immunosuppression or use within 16 weeks before randomization was an exclusion criterion. | 152 |
|  | Concurrent matching placebo | 43 [18–83]a | 40.5 | 63.36 (25.34) | 1.28 [0.5–5.46]a | RASi 97.0 SGLT2i 42.9 | Current systemic immunosuppression or use within 16 weeks before randomization was an exclusion criterion. | 168 |
| Lafayette 2024  ORIGIN phase IIb | Atacicept 150 mg  subcutaneously, once weekly | 38 (11) | 33.3 | 56 (23) | 1.4 [1–2.2] | RASi 97.0 SGLT2i 12.1 | Systemic treatment for IgAN within 3 months before screening or an anticipated need for such treatment was an exclusion criterion; exceptions applied to short-term use for other indications. | 33 |
|  | Concurrent matching placebo | 39 (13) | 44.1 | 66 (32) | 1.5 [0.9–2.1] | RASi 94.1 SGLT2i 17.6 | Systemic treatment for IgAN within 3 months before screening or an anticipated need for such treatment was an exclusion criterion; exceptions applied to short-term use for other indications. | 34 |
| Lafayette 2026  ORIGIN phase III | Atacicept 150 mg  subcutaneously, once weekly | 40.1 (11.1) | 46.2 | 65.3 (27.7) | 1.7 (0.9) | RASi 99.1 SGLT2i 55.7 | Pre-enrollment washout restrictions unclear. | 106 |
|  | Concurrent matching placebo | 40.9 (11.6) | 40.2 | 64.9 (29) | 1.8 (1.2) | RASi 100.0 SGLT2i 50.5 | Pre-enrollment washout restrictions unclear. | 97 |
| Lv 2026 TELIGAN | Telitacicept 240 mg  subcutaneously, once weekly | 37.8 (9.4) | 51.6 | 76.53 (26.95) | 1.23 [0.47–5.86]a | RASi not obtained;  SGLT2i 35.2 | Washout of systemic corticosteroids or other immunosuppression ≥12 weeks | 159 |
|  | Concurrent matching placebo | 38.6 (10) | 56.0 | 74.43 (27.2) | 1.29 [0.54–6.59]a | RASi not obtained;  SGLT2i 34.6 | Washout of systemic corticosteroids or other immunosuppression ≥12 weeks | 159 |
| **b Additional studies in the 24–52-week broad window (4 RCTs)** | | | | | | | | |
| Heerspink 2025 SC0062 | SC0062 20 mg  orally, once daily | 43 (11) | 56.2 | 70 (23) | 1.3 [1–1.6] | RASi 100.0 SGLT2i 46.9 | Systemic corticosteroids or other immunosuppression for >2 weeks within the preceding 3 months was an exclusion criterion. | 32 |
|  | Concurrent matching placebo | 43 (13) | 32.4 | 71 (24) | 1 [0.7–1.9] | RASi 100.0 SGLT2i 47.1 | Systemic corticosteroids or other immunosuppression for >2 weeks within the preceding 3 months was an exclusion criterion. | 34 |
| Lafayette 2025 SANCTUARY | Ravulizumab  intravenously, weight-based dosing | 41 (10) | 51.2 | 76 (30) | 1.4 [1.1–2.3] | ACEi 46.5；ARB 58.1c SGLT2i 20.9 | Systemic immunosuppression restricted within 6 months before screening | 43 |
|  | Concurrent matching placebo | 39 (12) | 34.8 | 70 (30) | 1.5 [1–2.7] | ACEi 30.4；ARB 69.6c SGLT2i 17.4 | Systemic immunosuppression restricted within 6 months before screening | 23 |
| Mathur 2024 ENVISION | Sibeprenlimab 4 mg/kg  intravenously, every 4 weeks | 39 [20–73]a | 36.6 | 64 [35–133]a | 1.53 (0.12)b | RASi 97.6 SGLT2i 4.9 | Systemic corticosteroids or other immunosuppression within 16 weeks before initial screening was an exclusion criterion | 41 |
|  | Concurrent matching placebo | 36 [18–52]a | 63.2 | 68.5 [33–116]a | 1.68 (0.17)b | RASi 100.0 SGLT2i 7.9 | Systemic corticosteroids or other immunosuppression within 16 weeks before initial screening was an exclusion criterion | 38 |
| Zhang 2024  Iptacopan phase II | Iptacopan 200 mg  orally, twice daily | Not obtained | Not obtained | Not obtained | Not obtained | RASi not obtained;  SGLT2i not obtained | Exposure to immunosuppressants or systemic corticosteroids within 90 days before dosing was an exclusion criterion | 11 |
|  | Concurrent matching placebo | Not obtained | Not obtained | Not obtained | Not obtained | RASi not obtained;  SGLT2i not obtained | Exposure to immunosuppressants or systemic corticosteroids within 90 days before dosing was an exclusion criterion | 11 |
| **c Studies not included in quantitative proteinuria analyses (4 RCTs)** | | | | | | | | |
| Lv 2023  Telitacicept phase II | Telitacicept 240 mg  subcutaneously, once weekly | 36.8 (8.83) | 64.3 | 70.17 [57.43–94.94] | Urinary protein¹ 1.34 [1.14–2.01]  g/d | RASi not obtained;  SGLT2i not obtained | Pre-enrollment washout restrictions unclear | 14 |
|  | Concurrent matching placebo | 38.3 (6.91) | 28.6 | 91.2 [67.73–108.83] | Urinary protein¹ 1.86 [1.57–2.36]  g/d | RASi not obtained;  SGLT2i not obtained | Pre-enrollment washout restrictions unclear | 14 |
| Floege 2025 IGNAZ | Felzartamab  intravenously, 9-dose regimen | 39.2 (7.2) | 23.1 | 70.7 (25.6) | 1.1 [0.6–2.7] | RASi 100.0 ; SGLT2i not obtained | Systemic immunosuppression for >7 consecutive days within 180 days before informed consent was an exclusion criterion | 13 |
|  | Concurrent matching placebo | 39.6 (11.8) | 25.0 | 77 (33.3) | 1.8 [1.3–2.9] | RASi 100.0 ; SGLT2i not obtained | Systemic immunosuppression for >7 consecutive days within 180 days before informed consent was an exclusion criterion | 12 |
| ClinicalTrials.gov 2025 ARTEMIS-IgAN | Narsoplimab 370 mg  intravenously | 40.7 (12.01) | 37.0 | Not obtained | 2 (1.21) | RASi not obtained;  SGLT2i not obtained | Pre-enrollment washout restrictions unclear | 181 d |
|  | Concurrent matching placebo | 41.6 (12.76) | 34.1 | Not obtained | 2.1 (1.19) | RASi not obtained;  SGLT2i not obtained | Pre-enrollment washout restrictions unclear | 179 |
| ClinicalTrials.gov 2025 ALXN2050 IgAN | Vemircopan 180 mg  orally, twice daily | Not obtained | 34.6 | Not obtained | Not obtained | RASi not obtained;  SGLT2i not obtained | Other systemic immunosuppression within the preceding 6 months was an exclusion criterion; prednisone-equivalent doses >20 mg/d for >14 days were also excluded | 26 |
|  | Concurrent matching placebo | Not obtained | 38.5 | Not obtained | Not obtained | RASi not obtained;  SGLT2i not obtained | Other systemic immunosuppression within the preceding 6 months was an exclusion criterion; prednisone-equivalent doses >20 mg/d for >14 days were also excluded | 26 |

Note: Values are means (SDs) or medians [Q1–Q3]; a indicates median [minimum–maximum]; b indicates geometric mean (GSE). ¹ The telitacicept phase II trial reported 24-hour urinary protein (g/d); all others reported 24-hour UPCR. Participant numbers per arm refer to the corresponding baseline populations; d indicates the registered number of participants who actually received treatment. RASi and SGLT2i represent actual baseline use; c indicates that the ACEi and ARB categories may overlap.

### **APPENDIX 5 Direct Evidence and Pairwise Analyses**

**Table 5.1 Direct evidence for proteinuria in the primary window (32–40 weeks)**

| **Comparison and study** | **Number analyzed, target /control** | **Effect measure** | **Estimate (95% CI)** | **τ²** | **I²（%）** | **Outcome time point** | **Overall RoB 2** |
| --- | --- | --- | --- | --- | --- | --- | --- |
| **Targeted-release budesonide 16 mg  vs placebo** | **462** | **GMR** | 0.70（0.62–0.79） | **0.000** | **0.0** |  |  |
| *Fellström 2017*  NEFIGAN | 98 (48/50) | GMR | 0.72（0.56–0.92） | Not applicable | Not applicable | UPCR: 9 months | Some concerns^2,3,5^ |
| *Lafayette 2023*  NefIgArd | 364 (182/182) | GMR | 0.70（0.61–0.80） | Not applicable | Not applicable | UPCR: 9 months; eGFR: time-weighted average over 24 months; total slope: 24 months. | Some concerns^3^ |
| **Atacicept 150 mg  vs placebo** | **270** | **GMR** | 0.60（0.51–0.71） | **0.000** | **0.0** |  |  |
| *Lafayette 2024*  ORIGIN phase IIb | 67 (33/34) | GMR | 0.65（0.47–0.91） | Not applicable | Not applicable | UPCR: 36 weeks | Some concerns^3^ |
| *Lafayette 2026*  ORIGIN phase III | 203 (106/97) | GMR | 0.58（0.48–0.71） | Not applicable | Not applicable | UPCR: 36 weeks. | Low risk |
| **Atrasentan 0.75 mg  vs placebo**  *Heerspink 2026*  ALIGN | 340 (170/170) | GMR | 0.62（0.54–0.70） | Not estimable | Not estimable | UPCR: 36 weeks; fixed-time-point eGFR: 136 weeks; total slope: baseline to 136 weeks. | Some concerns^3^ |
| **Iptacopan 200 mg  vs placebo**  *Barratt 2026*  APPLAUSE-IgAN | 477 (238/239) | GMR | 0.61（0.54–0.69） | Not estimable | Not estimable | Total eGFR slope: 24 months; UPCR: 9 months | Some concerns^3^ |
| **Sibeprenlimab 400 mg subcutaneously  vs placebo**  *Perkovic 2026*  VISIONARY | 320 (152/168) | GMR | 0.49（0.42–0.56） | Not estimable | Not estimable | UPCR: 9 months | Low risk |
| **Telitacicept 240 mg  vs placebo**  *Lv 2026*  TELIGAN | 318 (159/159) | GMR | 0.45（0.39–0.52） | Not estimable | Not estimable | UPCR: 39 weeks | Low risk |
| Cemdisiran 600 mg  vs placebo  Barratt 2024  Cemdisiran | 31 (22/9) | GMR | 0.63（0.36–1.10） | Not estimable | Not estimable | UPCR: 32 weeks | Some concerns³˒⁵ |

Pairwise synthesis: Bold comparison rows present inverse-variance common-effect estimates with normal-based 95% CIs. τ² was estimated separately by REML and is expressed in squared log(GMR) units; I² was calculated from Cochran’s Q. A zero estimate does not exclude clinically important heterogeneity.

**Table 5.2 Additional direct evidence for proteinuria in the broad window (24–52 weeks)**

| **Comparison and study** | **Number analyzed, target /control** | **Effect measure** | **Estimate (95% CI)** | **τ²** | **I²（%）** | **Outcome time point** | **Overall RoB 2** |
| --- | --- | --- | --- | --- | --- | --- | --- |
| **SC0062 20 mg  vs placebo**  *Heerspink 2025*  SC0062 | 66 (32/34) | GMR | 0.48（0.36–0.65） | Not estimable | Not estimable | 24 weeks | Some concerns^1,3^ |
| **Ravulizumab, weight-based dosing  vs placebo (1)**  *Lafayette 2025*  SANCTUARY | 66 (43/23) | GMR | 0.67（0.52–0.85） | Not estimable | Not estimable | 26 weeks | Low risk |
| **Sibeprenlimab 4 mg/kg intravenously  vs placebo**  *Mathur 2024*  ENVISION | 79 (41/38) | GMR | 0.52（0.35–0.77） | Not estimable | Not estimable | 12 months | Some concerns^3^ |
| **Iptacopan 200 mg twice daily  vs placebo**  *Zhang 2024*  Iptacopan phase II | 22 (11/11) | GMR | 0.80（0.46–1.39） | Not estimable | Not estimable | 180 days (25.7 weeks) | Some concerns³ |

**Table 5.3 Direct evidence for annualized total eGFR slope**

| **Comparison and study** | **Number analyzed, target /control** | **Effect measure** | **Estimate (95% CI)** | **τ²** | **I²（%）** | **Outcome time point and follow-up** | **Overall RoB 2** |
| --- | --- | --- | --- | --- | --- | --- | --- |
| **Targeted-release budesonide 16 mg  vs placebo**  *Lafayette 2023*  NefIgArd | 364 (182/182) | MD | 1.82（0.50–3.13） | Not estimable | Not estimable | 24 months | High risk^3^ |
| **Atrasentan 0.75 mg  vs placebo**  *Heerspink 2026*  ALIGN | 340 (170/170) | MD | 1.41（0.48–2.34） | Not estimable | Not estimable | 136 weeks | High risk^3^ |
| **Iptacopan 200 mg  vs placebo**  *Barratt 2026*  APPLAUSE-IgAN | 477 (238/239) | MD | 3.02（2.02–4.01） | Not estimable | Not estimable | 24 months | Some concerns^3^ |

*Each direct comparison was supported by only 1 RCT. Total slope includes the acute phase and subsequent changes and was not combined with chronic slope, fixed-time-point eGFR changes, or time-weighted average eGFR.*

**Table 5.4 Direct evidence for composite kidney outcomes**

| **Comparison and study** | **Number analyzed, target /control** | **Effect measure** | **Estimate (95% CI)** | **τ²** | **I²（%）** | **Outcome time point** | **Overall RoB 2** |
| --- | --- | --- | --- | --- | --- | --- | --- |
| **Targeted-release budesonide 16 mg  vs placebo**  *Lafayette 2023*  NefIgArd | 364 (182/182) | HR | 0.44（0.27–0.71） | Not estimable | Not estimable | 24 months | Some concerns^3,5^ |
| **Iptacopan 200 mg  vs placebo (1)**  *Barratt 2026*  APPLAUSE-IgAN | 477 (238/239) | HR | 0.57（0.40–0.81） | Not estimable | Not estimable | 24 months | Some concerns^3^ |

**Table 5.5 Direct evidence for serious adverse events (24–26 weeks)**

| **Comparison and study** | **Number analyzed, target /control** | **Effect measure** | **Estimate (95% CI)** | **τ²** | **I²（%）** | **Outcome time point** | **Overall RoB 2** |
| --- | --- | --- | --- | --- | --- | --- | --- |
| **SC0062 20 mg  vs placebo**  *Heerspink 2025*  SC0062 | 65 (31/34) SAE 2/31 vs 1/34 | OR | 1.89（0.23–15.70） | Not estimable | Not estimable | SAEs: 24 weeks of treatment and 4 weeks after the last dose | High risk^1,2,3^ |
| **Ravulizumab, weight-based dosing  vs placebo**  *Lafayette 2025*  SANCTUARY | 66 (43/23) SAE 1/43 vs 0/23 | OR | 1.66（0.06–45.05） | Not estimable | Not estimable | SAEs: 26 weeks | Some concerns^3^ |

Note: Event counts represent participants experiencing at least one serious adverse event. ORs and 95% CIs were obtained from the common-effect penalized-likelihood logistic regression (LRP) network model. The Mantel–Haenszel (MH) sensitivity analysis without continuity correction is presented in Table 8.5; the ravulizumab control arm had zero events, and MH did not yield a finite estimate. The finite LRP estimate incorporates information contributed by the penalty. The early-window network included only 2 RCTs, with residual df = 0; between-study heterogeneity could not be estimated independently.

**Table 5.6 Direct evidence for serious adverse events (36–39 weeks)**

| **Comparison and study** | **Number analyzed, target /control** | **Effect measure** | **Estimate (95% CI)** | **τ²** | **I²（%）** | **Outcome time point and follow-up** | **Overall RoB 2** |
| --- | --- | --- | --- | --- | --- | --- | --- |
| **Cemdisiran 600 mg  vs placebo**  *Barratt 2024*  Cemdisiran | 31 (22/9) SAE 1/22 vs 0/9 | OR | 1.33（0.04–41.44） | Not estimable | Not estimable | SAEs: 36-week randomized double-blind period | Some concerns^3^ |
| **Atacicept 150 mg  vs placebo**  *Lafayette 2024*  ORIGIN phase IIb | 67 (33/34) SAE 1/33 vs 3/34 | OR | 0.42（0.06–3.08） | Not estimable | Not estimable | SAEs: through week 36, randomized double-blind period | Some concerns^3^ |
| **Telitacicept 240 mg  vs placebo**  *Lv 2026*  TELIGAN | 318 (159/159) SAE 4/159 vs 13/159 | OR | 0.31（0.11–0.94） | Not estimable | Not estimable | SAEs: after the first dose to before dosing at week 40 | Some concerns^3^ |

Note: ORs and 95% CIs were obtained from the LRP common-effect network model.

**Table 5.7 Direct common-effect and network random-effects estimates for proteinuria in the primary window**

| **Regimen** | **No. of direct RCTs** | **Direct evidence: common-effect model** | **NMA: random-effects t intervals** |
| --- | --- | --- | --- |
| Atacicept 150 mg | 2 | 0.60（0.51–0.71） | 0.60（0.41–0.87） |
| Atrasentan 0.75 mg | 1 | 0.62（0.54–0.70） | 0.62（0.47–0.81） |
| Iptacopan 200 mg | 1 | 0.61（0.54–0.69） | 0.61（0.47–0.80） |
| Sibeprenlimab 400 mg (subcutaneous) | 1 | 0.49（0.42–0.56） | 0.49（0.35–0.67） |
| Targeted-release budesonide 16 mg | 2 | 0.70（0.62–0.79） | 0.70（0.54–0.91） |
| Telitacicept 240 mg | 1 | 0.45（0.39–0.52） | 0.45（0.32–0.63） |
| Cemdisiran 600 mg | 1 | 0.63（0.36–1.10） | 0.63（0.18–2.16） |

Note: Data are GMRs (95% CIs). Direct evidence was synthesized using an inverse-variance common-effect model; for single-study comparisons, intervals were calculated from the corresponding standard errors using the normal distribution. NMA used a random-effects model, with τ² estimated by REML and intervals constructed using the t distribution (df=2).

### **APPENDIX 6 Risk of bias and certainty of evidence**

#### **6.1 Result-specific risk-of-bias assessment**

**Table 6.1 RoB 2 assessments of proteinuria results**

| **Study and result assessed** | **D1  Randomization process** | **D2  Deviations from intended interventions** | **D3  Missing outcome data** | **D4  Measurement of the outcome** | **D5  Selection of the reported result** | **Overall risk of bias** |
| --- | --- | --- | --- | --- | --- | --- |
| **Fellström 2017 (NEFIGAN** ) Proteinuria · Week 39 | Low risk | Some concerns | Some concerns | Low risk | Some concerns | Some concerns |
| **Lafayette 2023 (NefIgArd** ) Proteinuria · Week 39 | Low risk | Low risk | Some concerns | Low risk | Low risk | Some concerns |
| **Heerspink 2026 (ALIGN** ) Proteinuria · Week 36 | Low risk | Low risk | Some concerns | Low risk | Low risk | Some concerns |
| **Heerspink 2025 (SC0062** ) Proteinuria · Week 24 | Some concerns | Low risk | Some concerns | Low risk | Low risk | Some concerns |
| **Lafayette 2025 (SANCTUARY** ) Proteinuria · Week 26 | Low risk | Low risk | Low risk | Low risk | Low risk | Low risk |
| **Barratt 2024 (Cemdisiran** ) Proteinuria · Week 32 | Low risk | Low risk | Some concerns | Low risk | Some concerns | Some concerns |
| **Barratt 2026 (APPLAUSE-IgAN** ) Proteinuria · Week 39 | Low risk | Low risk | Some concerns | Low risk | Low risk | Some concerns |
| **Mathur 2024 (ENVISION** ) Proteinuria · Week 52 | Low risk | Low risk | Some concerns | Low risk | Low risk | Some concerns |
| **Perkovic 2026 (VISIONARY** ) Proteinuria · Week 40 | Low risk | Low risk | Low risk | Low risk | Low risk | Low risk |
| **Lafayette 2024 (ORIGIN phase 2b** ) Proteinuria · Week 36 | Low risk | Low risk | Some concerns | Low risk | Low risk | Some concerns |
| **Lafayette 2026 (ORIGIN phase 3** ) Proteinuria · Week 36 | Low risk | Low risk | Low risk | Low risk | Low risk | Low risk |
| Lv 2026 (TELIGAN ) Proteinuria · Week 39 | Low risk | Low risk | Low risk | Low risk | Low risk | Low risk |
| **Zhang 2024 (Iptacopan)**  Proteinuria · Day 180 | Low risk | Low risk | Some concerns | Low risk | Low risk | Some concerns |

*Note: Green indicates low risk, yellow indicates some concerns, and red indicates high risk.*

**Table 6.2 RoB 2 assessments of kidney outcomes and serious adverse events**

| **Study and result assessed** | **D1  Randomization process** | **D2  Deviations from intended interventions** | **D3  Missing outcome data** | **D4  Measurement of the outcome** | **D5  Selection of the reported result** | **Overall risk of bias** |
| --- | --- | --- | --- | --- | --- | --- |
| **Lafayette 2023 (NefIgArd** ) Annualized total eGFR slope · Week 104 | Low risk | Low risk | High risk | Low risk | Low risk | High risk |
| **Heerspink 2026 (ALIGN** ) Annualized total eGFR slope · Week 136 | Low risk | Low risk | High risk | Low risk | Low risk | High risk |
| **Barratt 2026 (APPLAUSE-IgAN** ) Annualized total eGFR slope · Week 104 | Low risk | Low risk | Some concerns | Low risk | Low risk | Some concerns |
| **Lafayette 2023 (NefIgArd** ) Composite kidney outcome · Week 104 | Low risk | Low risk | Some concerns | Low risk | Some concerns | Some concerns |
| **Barratt 2026 (APPLAUSE-IgAN** ) Composite kidney outcome · Week 104 | Low risk | Low risk | Some concerns | Low risk | Low risk | Some concerns |
| **Heerspink 2025 (SC0062** ) Serious adverse events · 24 weeks of treatment and 4 weeks after the last dose | Some concerns | High risk | Some concerns | Low risk | Low risk | High risk |
| **Lafayette 2025 (SANCTUARY** ) Serious adverse events · Week 26 | Low risk | Low risk | Some concerns | Low risk | Low risk | Some concerns |
| **Barratt 2024 (Cemdisiran** ) Serious adverse events · Week 36 | Low risk | Low risk | Some concerns | Low risk | Low risk | Some concerns |
| **Lafayette 2024 (ORIGIN** ) Serious adverse events · Week 36 | Low risk | Low risk | Some concerns | Low risk | Low risk | Some concerns |
| **Lv 2026 (TELIGAN** ) Serious adverse events · Week 39 | Low risk | Low risk | Some concerns | Low risk | Low risk | Some concerns |

*Note: Green indicates low risk, yellow indicates some concerns, and red indicates high risk. RoB 2 assessments are result-specific; judgments for one outcome are not directly extrapolated to other outcomes from the same trial.*

#### **6.2 Bias due to missing evidence and certainty of evidence**

**Table 6.3 Summary of ROB-MEN and CINeMA assessments**

| **Analysis set** | **No. of comparisons** | **ROB-MEN  low risk/some concerns** | **CINeMA : Low certainty** | **CINeMA : Very low certainty** |
| --- | --- | --- | --- | --- |
| Proteinuria, primary window: 32–40 weeks | 28 | 7/21 | 3 | 25 |
| Proteinuria, broad window: 24–52 weeks | 55 | 10/45 | 4 | 51 |
| Annualized total eGFR slope | 6 | 3/3 | 0 | 6 |
| Composite kidney outcome | 3 | 2/1 | 0 | 3 |
| Serious adverse events: 24–26 weeks | 3 | 2/1 | 0 | 3 |
| Serious adverse events: 36–39 weeks | 6 | 3/3 | 0 | 6 |

Note: The number of comparisons includes comparisons of active treatments with the trial-defined control and indirect comparisons between active treatments.

**Table 6.4 CINeMA assessments for proteinuria in the primary window (32–40 weeks)**

| **Treatment comparison** | **Evidence** | **GMR （95% CI）** | **Within-study bias** | **Reporting bias** | **Indirectness** | **Imprecision** | **Heterogeneity** | **Incoherence** | **Overall certainty** |
| --- | --- | --- | --- | --- | --- | --- | --- | --- | --- |
| Atacicept 150 mg  vs trial-defined control | Direct | 0.599 (0.412–0.872) | No concerns | No concerns | No concerns | No concerns | Some concerns | Major concerns | Low |
| Atrasentan 0.75 mg  vs trial-defined control | Direct | 0.617 (0.468–0.814) | Some concerns | No concerns | No concerns | No concerns | Some concerns | Major concerns | Very low |
| Cemdisiran 600 mg  vs trial-defined control | Direct | 0.626 (0.182–2.156) | Some concerns | No concerns | No concerns | Major concerns | Some concerns | Major concerns | Very low |
| Iptacopan 200 mg twice  daily vs trial-defined control | Direct | 0.610 (0.466–0.798) | Some concerns | No concerns | No concerns | No concerns | Some concerns | Major concerns | Very low |
| Sibeprenlimab 400 mg subcutaneous  vs trial-defined control | Direct | 0.488 (0.355–0.671) | No concerns | No concerns | No concerns | No concerns | Some concerns | Major concerns | Low |
| Targeted-release budesonide 16 mg  vs trial-defined control | Direct | 0.704 (0.542–0.914) | Some concerns | No concerns | No concerns | No concerns | Some concerns | Major concerns | Very low |
| Telitacicept 240 mg  vs trial-defined control | Direct | 0.450 (0.323–0.628) | No concerns | No concerns | No concerns | No concerns | Some concerns | Major concerns | Low |
| Atacicept 150 mg  vs Atrasentan 0.75 mg | Indirect | 0.972 (0.610–1.548) | Some concerns | Some concerns | Some concerns | Major concerns | Some concerns | Major concerns | Very low |
| Atacicept 150 mg  vs Cemdisiran 600 mg | Indirect | 0.957 (0.263–3.482) | Some concerns | Some concerns | Some concerns | Major concerns | Some concerns | Major concerns | Very low |
| Atacicept 150 mg  vs Iptacopan 200 mg twice daily | Indirect | 0.983 (0.619–1.559) | Some concerns | Some concerns | Some concerns | Major concerns | Some concerns | Major concerns | Very low |
| Atacicept 150 mg  vs Sibeprenlimab 400 mg subcutaneous | Indirect | 1.228 (0.751–2.009) | No concerns | Some concerns | Some concerns | Major concerns | Some concerns | Major concerns | Very low |
| Atacicept 150 mg  vs Targeted-release budesonide 16 mg | Indirect | 0.852 (0.539–1.345) | Some concerns | Some concerns | Some concerns | Major concerns | Some concerns | Major concerns | Very low |
| Atacicept 150 mg  vs Telitacicept 240 mg | Indirect | 1.332 (0.807–2.199) | No concerns | Some concerns | Some concerns | Some concerns | Some concerns | Major concerns | Very low |
| Atrasentan 0.75 mg  vs Cemdisiran 600 mg | Indirect | 0.985 (0.278–3.496) | Some concerns | Some concerns | Some concerns | Major concerns | Some concerns | Major concerns | Very low |
| Atrasentan 0.75 mg  vs Iptacopan 200 mg twice daily | Indirect | 1.011 (0.688–1.488) | Some concerns | Some concerns | Some concerns | Major concerns | Some concerns | Major concerns | Very low |
| Atrasentan 0.75 mg  vs Sibeprenlimab 400 mg subcutaneous | Indirect | 1.264 (0.829–1.928) | Some concerns | Some concerns | Some concerns | Some concerns | Some concerns | Major concerns | Very low |
| Atrasentan 0.75 mg  vs Targeted-release budesonide 16 mg | Indirect | 0.877 (0.599–1.283) | Some concerns | Some concerns | Some concerns | Major concerns | Some concerns | Major concerns | Very low |
| Atrasentan 0.75 mg  vs Telitacicept 240 mg | Indirect | 1.371 (0.889–2.114) | Some concerns | Some concerns | Some concerns | Some concerns | Some concerns | Major concerns | Very low |
| Cemdisiran 600 mg  vs Iptacopan 200 mg twice daily | Indirect | 1.027 (0.290–3.637) | Some concerns | Some concerns | Some concerns | Major concerns | Some concerns | Major concerns | Very low |
| Cemdisiran 600 mg  vs Sibeprenlimab 400 mg subcutaneous | Indirect | 1.283 (0.358–4.599) | Some concerns | Some concerns | Some concerns | Major concerns | Some concerns | Major concerns | Very low |
| Cemdisiran 600 mg  vs Targeted-release budesonide 16 mg | Indirect | 0.890 (0.252–3.148) | Some concerns | Some concerns | Some concerns | Major concerns | Some concerns | Major concerns | Very low |
| Cemdisiran 600 mg  vs Telitacicept 240 mg | Indirect | 1.392 (0.387–5.005) | Some concerns | Some concerns | Some concerns | Major concerns | Some concerns | Major concerns | Very low |
| Iptacopan 200 mg twice daily vs  Sibeprenlimab 400 mg subcutaneous | Indirect | 1.250 (0.824–1.896) | Some concerns | Some concerns | Some concerns | Some concerns | Some concerns | Major concerns | Very low |
| Iptacopan 200 mg twice  daily vs Targeted-release budesonide 16 mg | Indirect | 0.867 (0.596–1.261) | Some concerns | Some concerns | Some concerns | Major concerns | Some concerns | Major concerns | Very low |
| Iptacopan 200 mg twice daily vs  Telitacicept 240 mg | Indirect | 1.356 (0.884–2.079) | Some concerns | Some concerns | Some concerns | Some concerns | Some concerns | Major concerns | Very low |
| Sibeprenlimab 400 mg subcutaneous  vs Targeted-release budesonide 16 mg | Indirect | 0.693 (0.459–1.047) | Some concerns | Some concerns | Some concerns | Some concerns | Some concerns | Major concerns | Very low |
| Sibeprenlimab 400 mg subcutaneous vs  Telitacicept 240 mg | Indirect | 1.084 (0.684–1.718) | No concerns | Some concerns | Some concerns | Major concerns | Some concerns | Major concerns | Very low |
| Targeted-release budesonide 16 mg  vs Telitacicept 240 mg | Indirect | 1.564 (1.025–2.387) | Some concerns | Some concerns | Some concerns | No concerns | Some concerns | Major concerns | Very low |

**Table 6.5 CINeMA assessments for proteinuria in the broad window (24–52 weeks)**

| **Treatment comparison** | **Evidence** | **GMR （95% CI）** | **Within-study bias** | **Reporting bias** | **Indirectness** | **Imprecision** | **Heterogeneity** | **Incoherence** | **Overall certainty** |
| --- | --- | --- | --- | --- | --- | --- | --- | --- | --- |
| Atacicept 150 mg  vs trial-defined control | Direct | 0.599 (0.454–0.791) | No concerns | No concerns | No concerns | No concerns | Some concerns | Major concerns | Low |
| Atrasentan 0.75 mg  vs trial-defined control | Direct | 0.617 (0.503–0.757) | Some concerns | No concerns | No concerns | No concerns | Some concerns | Major concerns | Very low |
| Cemdisiran 600 mg  vs trial-defined control | Direct | 0.626 (0.251–1.562) | Some concerns | No concerns | No concerns | Major concerns | Some concerns | Major concerns | Very low |
| Iptacopan 200 mg twice  daily vs trial-defined control | Direct | 0.618 (0.509–0.750) | Some concerns | No concerns | Some concerns | Some concerns | Some concerns | Major concerns | Very low |
| Ravulizumab, weight-based  dosing vs trial-defined control | Direct | 0.668 (0.450–0.991) | No concerns | No concerns | No concerns | No concerns | Some concerns | Major concerns | Low |
| SC0062 20 mg  vs trial-defined control | Direct | 0.484 (0.297–0.789) | Some concerns | No concerns | No concerns | No concerns | Some concerns | Major concerns | Very low |
| Sibeprenlimab 400 mg subcutaneous  vs trial-defined control | Direct | 0.488 (0.386–0.618) | No concerns | No concerns | No concerns | No concerns | Some concerns | Major concerns | Low |
| Sibeprenlimab 4 mg/kg intravenous  vs trial-defined control | Direct | 0.516 (0.271–0.980) | Some concerns | No concerns | No concerns | No concerns | Some concerns | Major concerns | Very low |
| Targeted-release budesonide 16 mg  vs trial-defined control | Direct | 0.704 (0.580–0.854) | Some concerns | No concerns | No concerns | No concerns | Some concerns | Major concerns | Very low |
| Telitacicept 240 mg  vs trial-defined control | Direct | 0.450 (0.352–0.576) | No concerns | No concerns | No concerns | No concerns | Some concerns | Major concerns | Low |
| Atacicept 150 mg  vs Atrasentan 0.75 mg | Indirect | 0.972 (0.688–1.371) | Some concerns | Some concerns | Some concerns | Major concerns | Some concerns | Major concerns | Very low |
| Atacicept 150 mg  vs Cemdisiran 600 mg | Indirect | 0.957 (0.368–2.488) | Some concerns | Some concerns | Some concerns | Major concerns | Some concerns | Major concerns | Very low |
| Atacicept 150 mg  vs Iptacopan 200 mg twice daily | Indirect | 0.970 (0.691–1.361) | Some concerns | Some concerns | Some concerns | Major concerns | Some concerns | Major concerns | Very low |
| Atacicept 150 mg  vs Ravulizumab, weight-based dosing | Indirect | 0.897 (0.554–1.453) | No concerns | Some concerns | Some concerns | Major concerns | Some concerns | Major concerns | Very low |
| Atacicept 150 mg  vs SC0062 20 mg | Indirect | 1.238 (0.706–2.173) | Some concerns | Some concerns | Some concerns | Major concerns | Some concerns | Major concerns | Very low |
| Atacicept 150 mg  vs Sibeprenlimab 400 mg subcutaneous | Indirect | 1.228 (0.854–1.767) | No concerns | Some concerns | Some concerns | Some concerns | Some concerns | Major concerns | Very low |
| Atacicept 150 mg  vs Sibeprenlimab 4 mg/kg intravenous | Indirect | 1.163 (0.578–2.340) | Some concerns | Some concerns | Some concerns | Major concerns | Some concerns | Major concerns | Very low |
| Atacicept 150 mg  vs Targeted-release budesonide 16 mg | Indirect | 0.852 (0.608–1.194) | Some concerns | Some concerns | Some concerns | Some concerns | Some concerns | Major concerns | Very low |
| Atacicept 150 mg  vs Telitacicept 240 mg | Indirect | 1.332 (0.919–1.930) | No concerns | Some concerns | Some concerns | Some concerns | Some concerns | Major concerns | Very low |
| Atrasentan 0.75 mg  vs Cemdisiran 600 mg | Indirect | 0.985 (0.386–2.514) | Some concerns | Some concerns | Some concerns | Major concerns | Some concerns | Major concerns | Very low |
| Atrasentan 0.75 mg  vs Iptacopan 200 mg twice daily | Indirect | 0.999 (0.753–1.324) | Some concerns | Some concerns | Some concerns | Major concerns | Some concerns | Major concerns | Very low |
| Atrasentan 0.75 mg  vs Ravulizumab, weight-based dosing | Indirect | 0.924 (0.592–1.440) | Some concerns | Some concerns | Some concerns | Major concerns | Some concerns | Major concerns | Very low |
| Atrasentan 0.75 mg  vs SC0062 20 mg | Indirect | 1.275 (0.750–2.166) | Some concerns | Some concerns | Some concerns | Major concerns | Some concerns | Major concerns | Very low |
| Atrasentan 0.75 mg  vs Sibeprenlimab 400 mg subcutaneous | Indirect | 1.264 (0.926–1.727) | Some concerns | Some concerns | Some concerns | Some concerns | Some concerns | Major concerns | Very low |
| Atrasentan 0.75 mg  vs Sibeprenlimab 4 mg/kg intravenous | Indirect | 1.197 (0.610–2.348) | Some concerns | Some concerns | Some concerns | Major concerns | Some concerns | Major concerns | Very low |
| Atrasentan 0.75 mg  vs Targeted-release budesonide 16 mg | Indirect | 0.877 (0.662–1.162) | Some concerns | Some concerns | Some concerns | Some concerns | Some concerns | Major concerns | Very low |
| Atrasentan 0.75 mg  vs Telitacicept 240 mg | Indirect | 1.371 (0.996–1.888) | Some concerns | Some concerns | Some concerns | Some concerns | Some concerns | Major concerns | Very low |
| Cemdisiran 600 mg  vs Iptacopan 200 mg twice daily | Indirect | 1.014 (0.398–2.581) | Some concerns | Some concerns | Some concerns | Major concerns | Some concerns | Major concerns | Very low |
| Cemdisiran 600 mg  vs Ravulizumab, weight-based dosing | Indirect | 0.938 (0.347–2.537) | Some concerns | Some concerns | Some concerns | Major concerns | Some concerns | Major concerns | Very low |
| Cemdisiran 600 mg  vs SC0062 20 mg | Indirect | 1.294 (0.459–3.650) | Some concerns | Some concerns | Some concerns | Major concerns | Some concerns | Major concerns | Very low |
| Cemdisiran 600 mg  vs Sibeprenlimab 400 mg subcutaneous | Indirect | 1.283 (0.499–3.299) | Some concerns | Some concerns | Some concerns | Major concerns | Some concerns | Major concerns | Very low |
| Cemdisiran 600 mg  vs Sibeprenlimab 4 mg/kg intravenous | Indirect | 1.215 (0.398–3.713) | Some concerns | Some concerns | Some concerns | Major concerns | Some concerns | Major concerns | Very low |
| Cemdisiran 600 mg  vs Targeted-release budesonide 16 mg | Indirect | 0.890 (0.350–2.266) | Some concerns | Some concerns | Some concerns | Major concerns | Some concerns | Major concerns | Very low |
| Cemdisiran 600 mg  vs Telitacicept 240 mg | Indirect | 1.392 (0.540–3.587) | Some concerns | Some concerns | Some concerns | Major concerns | Some concerns | Major concerns | Very low |
| Iptacopan 200 mg twice daily vs  Ravulizumab, weight-based dosing | Indirect | 0.925 (0.596–1.435) | Some concerns | Some concerns | Some concerns | Major concerns | Some concerns | Major concerns | Very low |
| Iptacopan 200 mg twice daily vs  SC0062 20 mg | Indirect | 1.277 (0.754–2.161) | Some concerns | Some concerns | Some concerns | Major concerns | Some concerns | Major concerns | Very low |
| Iptacopan 200 mg twice daily vs  Sibeprenlimab 400 mg subcutaneous | Indirect | 1.266 (0.933–1.718) | Some concerns | Some concerns | Some concerns | Some concerns | Some concerns | Major concerns | Very low |
| Iptacopan 200 mg twice daily vs  Sibeprenlimab 4 mg/kg intravenous | Indirect | 1.199 (0.613–2.344) | Some concerns | Some concerns | Some concerns | Major concerns | Some concerns | Major concerns | Very low |
| Iptacopan 200 mg twice  daily vs Targeted-release budesonide 16 mg | Indirect | 0.878 (0.668–1.155) | Some concerns | Some concerns | Some concerns | Some concerns | Some concerns | Major concerns | Very low |
| Iptacopan 200 mg twice daily vs  Telitacicept 240 mg | Indirect | 1.373 (1.004–1.879) | Some concerns | Some concerns | Some concerns | Some concerns | Some concerns | Major concerns | Very low |
| Ravulizumab, weight-based dosing vs  SC0062 20 mg | Indirect | 1.380 (0.736–2.587) | Some concerns | Some concerns | Some concerns | Major concerns | Some concerns | Major concerns | Very low |
| Ravulizumab, weight-based dosing vs  Sibeprenlimab 400 mg subcutaneous | Indirect | 1.369 (0.865–2.166) | No concerns | Some concerns | Some concerns | Some concerns | Some concerns | Major concerns | Very low |
| Ravulizumab, weight-based dosing vs  Sibeprenlimab 4 mg/kg intravenous | Indirect | 1.296 (0.610–2.752) | Some concerns | Some concerns | Some concerns | Major concerns | Some concerns | Major concerns | Very low |
| Ravulizumab, weight-based  dosing vs Targeted-release budesonide 16 mg | Indirect | 0.949 (0.612–1.472) | Some concerns | Some concerns | Some concerns | Major concerns | Some concerns | Major concerns | Very low |
| Ravulizumab, weight-based dosing vs  Telitacicept 240 mg | Indirect | 1.484 (0.933–2.362) | No concerns | Some concerns | Some concerns | Some concerns | Some concerns | Major concerns | Very low |
| SC0062 20 mg  vs Sibeprenlimab 400 mg subcutaneous | Indirect | 0.992 (0.576–1.707) | Some concerns | Some concerns | Some concerns | Major concerns | Some concerns | Major concerns | Very low |
| SC0062 20 mg  vs Sibeprenlimab 4 mg/kg intravenous | Indirect | 0.939 (0.419–2.105) | Some concerns | Some concerns | Some concerns | Major concerns | Some concerns | Major concerns | Very low |
| SC0062 20 mg  vs Targeted-release budesonide 16 mg | Indirect | 0.688 (0.406–1.164) | Some concerns | Some concerns | Some concerns | Some concerns | Some concerns | Major concerns | Very low |
| SC0062 20 mg  vs Telitacicept 240 mg | Indirect | 1.076 (0.622–1.860) | Some concerns | Some concerns | Some concerns | Major concerns | Some concerns | Major concerns | Very low |
| Sibeprenlimab 400 mg subcutaneous vs  Sibeprenlimab 4 mg/kg intravenous | Indirect | 0.947 (0.478–1.876) | Some concerns | Some concerns | Some concerns | Major concerns | Some concerns | Major concerns | Very low |
| Sibeprenlimab 400 mg subcutaneous  vs Targeted-release budesonide 16 mg | Indirect | 0.693 (0.511–0.940) | Some concerns | Some concerns | Some concerns | No concerns | Some concerns | Major concerns | Very low |
| Sibeprenlimab 400 mg subcutaneous vs  Telitacicept 240 mg | Indirect | 1.084 (0.771–1.524) | No concerns | Some concerns | Some concerns | Major concerns | Some concerns | Major concerns | Very low |
| Sibeprenlimab 4 mg/kg intravenous  vs Targeted-release budesonide 16 mg | Indirect | 0.733 (0.375–1.432) | Some concerns | Some concerns | Some concerns | Major concerns | Some concerns | Major concerns | Very low |
| Sibeprenlimab 4 mg/kg intravenous vs  Telitacicept 240 mg | Indirect | 1.146 (0.576–2.278) | Some concerns | Some concerns | Some concerns | Major concerns | Some concerns | Major concerns | Very low |
| Targeted-release budesonide 16 mg  vs Telitacicept 240 mg | Indirect | 1.564 (1.144–2.138) | Some concerns | Some concerns | Some concerns | No concerns | Some concerns | Major concerns | Very low |

**Table 6.6 CINeMA assessments for the annualized total eGFR slope**

| **Treatment comparison** | **Evidence** | **MD （95% CI）** | **Within-study bias** | **Reporting bias** | **Indirectness** | **Imprecision** | **Heterogeneity** | **Incoherence** | **Overall certainty** |
| --- | --- | --- | --- | --- | --- | --- | --- | --- | --- |
| Atrasentan 0.75 mg  vs trial-defined control | Direct | 1.410 (0.489–2.331) | Major concerns | No concerns | No concerns | No concerns | Some concerns | Major concerns | Very low |
| Iptacopan 200 mg twice  daily vs trial-defined control | Direct | 3.020 (2.025–4.015) | Some concerns | No concerns | No concerns | No concerns | Some concerns | Major concerns | Very low |
| Targeted-release budesonide 16 mg  vs trial-defined control | Direct | 1.820 (0.505–3.135) | Major concerns | No concerns | No concerns | No concerns | Some concerns | Major concerns | Very low |
| Atrasentan 0.75 mg  vs Iptacopan 200 mg twice daily | Indirect | -1.610 (-2.966–-0.254) | Major concerns | Some concerns | Some concerns | No concerns | Some concerns | Major concerns | Very low |
| Atrasentan 0.75 mg  vs Targeted-release budesonide 16 mg | Indirect | -0.410 (-2.016–1.196) | Major concerns | Some concerns | Some concerns | Major concerns | Some concerns | Major concerns | Very low |
| Iptacopan 200 mg twice  daily vs Targeted-release budesonide 16 mg | Indirect | 1.200 (-0.449–2.849) | Major concerns | Some concerns | Some concerns | Some concerns | Some concerns | Major concerns | Very low |

**Table 6.7 CINeMA assessments for the composite kidney outcome**

| **Treatment comparison** | **Evidence** | **HR （95% CI）** | **Within-study bias** | **Reporting bias** | **Indirectness** | **Imprecision** | **Heterogeneity** | **Incoherence** | **Overall certainty** |
| --- | --- | --- | --- | --- | --- | --- | --- | --- | --- |
| Iptacopan 200 mg twice  daily vs trial-defined control | Direct | 0.570 (0.401–0.811) | Some concerns | No concerns | No concerns | No concerns | Some concerns | Major concerns | Very low |
| Targeted-release budesonide 16 mg  vs trial-defined control | Direct | 0.440 (0.271–0.714) | Some concerns | No concerns | No concerns | No concerns | Some concerns | Major concerns | Very low |
| Iptacopan 200 mg twice  daily vs Targeted-release budesonide 16 mg | Indirect | 1.295 (0.712–2.357) | Some concerns | Some concerns | Some concerns | Major concerns | Some concerns | Major concerns | Very low |

**Table 6.8 CINeMA assessments for serious adverse events (24–26 weeks)**

| **Treatment comparison** | **Evidence** | **OR （95% CI）** | **Within-study bias** | **Reporting bias** | **Indirectness** | **Imprecision** | **Heterogeneity** | **Incoherence** | **Overall certainty** |
| --- | --- | --- | --- | --- | --- | --- | --- | --- | --- |
| Ravulizumab, weight-based  dosing vs trial-defined control | Direct | 1.659 (0.061–45.047) | Some concerns | No concerns | No concerns | Major concerns | Some concerns | Major concerns | Very low |
| SC0062 20 mg  vs trial-defined control | Direct | 1.893 (0.228–15.696) | Major concerns | No concerns | No concerns | Major concerns | Some concerns | Major concerns | Very low |
| Ravulizumab, weight-based dosing vs  SC0062 20 mg | Indirect | 0.876 (0.017–44.225) | Major concerns | Some concerns | Some concerns | Major concerns | Some concerns | Major concerns | Very low |

**Table 6.9 CINeMA assessments for serious adverse events (36–39 weeks)**

| **Treatment comparison** | **Evidence** | **OR （95% CI）** | **Within-study bias** | **Reporting bias** | **Indirectness** | **Imprecision** | **Heterogeneity** | **Incoherence** | **Overall certainty** |
| --- | --- | --- | --- | --- | --- | --- | --- | --- | --- |
| Atacicept 150 mg  vs trial-defined control | Direct | 0.415 (0.056–3.081) | Some concerns | No concerns | No concerns | Major concerns | Some concerns | Major concerns | Very low |
| Cemdisiran 600 mg  vs trial-defined control | Direct | 1.326 (0.042–41.440) | Some concerns | No concerns | No concerns | Major concerns | Some concerns | Major concerns | Very low |
| Telitacicept 240 mg  vs trial-defined control | Direct | 0.314 (0.105–0.938) | Some concerns | No concerns | No concerns | No concerns | Some concerns | Major concerns | Very low |
| Atacicept 150 mg  vs Cemdisiran 600 mg | Indirect | 0.313 (0.006–16.824) | Some concerns | Some concerns | Some concerns | Major concerns | Some concerns | Major concerns | Very low |
| Atacicept 150 mg  vs Telitacicept 240 mg | Indirect | 1.323 (0.135–12.971) | Some concerns | Some concerns | Some concerns | Major concerns | Some concerns | Major concerns | Very low |
| Cemdisiran 600 mg  vs Telitacicept 240 mg | Indirect | 4.221 (0.114–156.350) | Some concerns | Some concerns | Some concerns | Major concerns | Some concerns | Major concerns | Very low |

### **APPENDIX 7 Network meta-analysis results**

#### **7.1 Network size and model specifications**

**Table 7.1 Network size and model specifications for each analysis set**

| **Analysis set** | **Outcome and time window** | **No. of RCTs** | **Analysis set N** | **No. of nodes** | **No. of direct edges** | **Residual df** | **Model** | **Effect scale** | **Interval method** |
| --- | --- | --- | --- | --- | --- | --- | --- | --- | --- |
| Proteinuria, primary window | 24-hour UPCR , 32–40 weeks | 9 | 2,218 | 8 | 7 | 2 | Random effects (REML) | GMR  fitted on the log scale | Primary model: t distribution, df=2 |
| Proteinuria, broad window | 24-hour UPCR , 24–52 weeks | 13 | 2,451 | 11 | 10 | 3 | Random effects (REML) | GMR  fitted on the log scale | Primary model: t distribution, df=3 |
| Exploratory total eGFR slope analysis | Total slope over  104 or 136 weeks | 3 | 1,181 | 4 | 3 | 0 | Inverse-variance common-effect model | MD , mL/min/1.73 m²/year | Conditional normal-based 95% intervals |
| Exploratory composite kidney outcome analysis | Composite kidney outcome at  104 weeks | 2 | 841 | 3 | 2 | 0 | Inverse-variance common-effect model | HR  fitted on the log scale | Conditional normal-based 95% intervals |
| Exploratory early-window SAE analysis | At least one SAE , 24–26 weeks | 2 | 131 | 3 | 2 | 0 | LRP penalized-likelihood common-effect model; MH sensitivity analysis | OR | Normal Wald 95% intervals for the LRP model; MH without continuity correction |
| Exploratory late-window SAE analysis | At least one SAE , 36–39 weeks (approximate risk window) | 3 | 416 | 4 | 3 | 0 | LRP penalized-likelihood common-effect model; MH sensitivity analysis | OR | Normal Wald 95% intervals for the LRP model; MH without continuity correction |

Note: Nodes include the trial-defined control; none of the networks contains a closed loop. Proteinuria analyses used REML random-effects models and t intervals based on the network residual degrees of freedom, without the full Hartung–Knapp variance adjustment. Other outcomes used common-effect models; heterogeneity cannot be independently estimated when the residual df=0. LRP denotes the penalized-likelihood model; MH denotes the Mantel–Haenszel sensitivity analysis without continuity correction. Effect direction: GMRs, HRs, and ORs below 1 favor the first-listed treatment; eGFR MDs above 0 favor the first-listed treatment. Comparisons between active treatments are based entirely on indirect evidence.

**7.2 Relative Effects of Targeted Therapies and CINeMA Certainty of Evidence**

**Table 7.2 Proteinuria primary window, 32–40 weeks**

| **GMR（95% CI）** | **Placebo** | **Atacicept 150 mg** | **Atrasentan 0.75 mg** | **Cemdisiran 600 mg** | **Iptacopan 200 mg** | **Sibeprenlimab 400 mg SC** | **Budesonide 16 mg TRF** | **Telitacicept 240 mg** |
| --- | --- | --- | --- | --- | --- | --- | --- | --- |
| **Placebo** | 1.00 |  |  |  |  |  |  |  |
| **Atacicept 150 mg** | 0.599^**^ (0.412, 0.872) | 1.00 |  |  |  |  |  |  |
| **Atrasentan 0.75 mg** | 0.617^*^ (0.468, 0.814) | 1.029^*^ (0.646, 1.641) | 1.00 |  |  |  |  |  |
| **Cemdisiran 600 mg** | 0.626^*^ (0.182, 2.156) | 1.045^*^ (0.287, 3.802) | 1.015^*^ (0.286, 3.602) | 1.00 |  |  |  |  |
| **Iptacopan 200 mg** | 0.610^*^ (0.466, 0.798) | 1.018^*^ (0.641, 1.615) | 0.989^*^ (0.672, 1.454) | 0.974^*^ (0.275, 3.450) | 1.00 |  |  |  |
| **Sibeprenlimab 400 mg SC** | 0.488^**^ (0.355, 0.671) | 0.814^*^ (0.498, 1.331) | 0.791^*^ (0.519, 1.206) | 0.779^*^ (0.217, 2.792) | 0.800^*^ (0.527, 1.214) | 1.00 |  |  |
| **Budesonide 16 mg TRF** | 0.704^*^ (0.542, 0.914) | 1.174^*^ (0.743, 1.854) | 1.141^*^ (0.780, 1.668) | 1.124^*^ (0.318, 3.973) | 1.154^*^ (0.793, 1.678) | 1.442^*^ (0.955, 2.176) | 1.00 |  |
| **Telitacicept 240 mg** | 0.450^**^ (0.323, 0.628) | 0.751^*^ (0.455, 1.239) | 0.729^*^ (0.473, 1.124) | 0.718^*^ (0.200, 2.584) | 0.738^*^ (0.481, 1.132) | 0.922^*^ (0.582, 1.461) | 0.639^*^ (0.419, 0.976) | 1.00 |

**Table 7.3 Proteinuria broad window, 24–52 weeks**

| **GMR（95% CI）** | **Placebo** | **Atacicept 150 mg** | **Atrasentan 0.75 mg** | **Cemdisiran 600 mg** | **Iptacopan 200 mg** | **Ravulizumab , weight-based dosing** | **SC0062 20 mg** | **Sibeprenlimab 400 mg SC** | **Sibeprenlimab 4 mg/kg IV** | **Budesonide 16 mg TRF** | **Telitacicept 240 mg** |
| --- | --- | --- | --- | --- | --- | --- | --- | --- | --- | --- | --- |
| **Placebo** | 1.00 |  |  |  |  |  |  |  |  |  |  |
| **Atacicept 150 mg** | 0.599^**^ (0.454, 0.791) | 1.00 |  |  |  |  |  |  |  |  |  |
| **Atrasentan 0.75 mg** | 0.617^*^ (0.503, 0.757) | 1.029^*^ (0.729, 1.453) | 1.00 |  |  |  |  |  |  |  |  |
| **Cemdisiran 600 mg** | 0.626^*^ (0.251, 1.562) | 1.045^*^ (0.402, 2.716) | 1.015^*^ (0.398, 2.590) | 1.00 |  |  |  |  |  |  |  |
| **Iptacopan 200 mg** | 0.618^*^ (0.509, 0.750) | 1.031^*^ (0.735, 1.446) | 1.001^*^ (0.755, 1.328) | 0.986^*^ (0.387, 2.512) | 1.00 |  |  |  |  |  |  |
| **Ravulizumab , weight-based dosing** | 0.668^**^ (0.450, 0.991) | 1.114^*^ (0.688, 1.804) | 1.083^*^ (0.694, 1.688) | 1.067^*^ (0.394, 2.886) | 1.081^*^ (0.697, 1.678) | 1.00 |  |  |  |  |  |
| **SC0062 20 mg** | 0.484^*^ (0.297, 0.789) | 0.807^*^ (0.460, 1.417) | 0.784^*^ (0.462, 1.333) | 0.773^*^ (0.274, 2.179) | 0.783^*^ (0.463, 1.326) | 0.725^*^ (0.387, 1.358) | 1.00 |  |  |  |  |
| **Sibeprenlimab 400 mg SC** | 0.488^**^ (0.386, 0.618) | 0.814^*^ (0.566, 1.171) | 0.791^*^ (0.579, 1.080) | 0.779^*^ (0.303, 2.002) | 0.790^*^ (0.582, 1.072) | 0.731^*^ (0.462, 1.156) | 1.008^*^ (0.586, 1.735) | 1.00 |  |  |  |
| **Sibeprenlimab 4 mg/kg IV** | 0.516^*^ (0.271, 0.980) | 0.860^*^ (0.427, 1.731) | 0.835^*^ (0.426, 1.639) | 0.823^*^ (0.269, 2.515) | 0.834^*^ (0.427, 1.632) | 0.772^*^ (0.363, 1.639) | 1.065^*^ (0.475, 2.387) | 1.056^*^ (0.533, 2.093) | 1.00 |  |  |
| **Budesonide 16 mg TRF** | 0.704^*^ (0.580, 0.854) | 1.174^*^ (0.837, 1.646) | 1.141^*^ (0.861, 1.511) | 1.124^*^ (0.441, 2.860) | 1.139^*^ (0.866, 1.498) | 1.053^*^ (0.679, 1.634) | 1.454^*^ (0.859, 2.460) | 1.442^*^ (1.064, 1.955) | 1.365^*^ (0.698, 2.669) | 1.00 |  |
| **Telitacicept 240 mg** | 0.450^**^ (0.352, 0.576) | 0.751^*^ (0.518, 1.088) | 0.729^*^ (0.530, 1.004) | 0.718^*^ (0.279, 1.852) | 0.728^*^ (0.532, 0.996) | 0.674^*^ (0.423, 1.072) | 0.930^*^ (0.538, 1.608) | 0.922^*^ (0.656, 1.296) | 0.873^*^ (0.439, 1.736) | 0.639^*^ (0.468, 0.874) | 1.00 |

**Table 7.4 Annualized total eGFR slope**

| **MD（95% CI）** | **Placebo** | **Atrasentan 0.75 mg** | **Iptacopan 200 mg** | **Budesonide 16 mg TRF** |
| --- | --- | --- | --- | --- |
| **Placebo** | 0.00 |  |  |  |
| **Atrasentan 0.75 mg** | 1.410^*^ (0.489, 2.331) | 0.00 |  |  |
| **Iptacopan 200 mg** | 3.020^*^ (2.025, 4.015) | 1.610^*^ (0.254, 2.966) | 0.00 |  |
| **Budesonide 16 mg TRF** | 1.820^*^ (0.505, 3.135) | 0.410^*^ (−1.196, 2.016) | −1.200^*^ (−2.849, 0.449) | 0.00 |

**Table 7.5 Composite kidney outcome, 104 weeks**

| **HR（95% CI）** | **Placebo** | **Iptacopan 200 mg** | **Budesonide 16 mg TRF** |
| --- | --- | --- | --- |
| **Placebo** | 1.00 |  |  |
| **Iptacopan 200 mg** | 0.570^*^ (0.401, 0.811) | 1.00 |  |
| **Budesonide 16 mg TRF** | 0.440^*^ (0.271, 0.714) | 0.772^*^ (0.424, 1.404) | 1.00 |

**Table 7.6 Serious adverse events, 24–26-week risk window**

| **OR（95% CI）** | **Placebo** | **Ravulizumab , weight-based dosing** | **SC0062 20 mg** |
| --- | --- | --- | --- |
| **Placebo** | 1.00 |  |  |
| **Ravulizumab , weight-based dosing** | 1.659^*^ (0.061, 45.047) | 1.00 |  |
| **SC0062 20 mg** | 1.893^*^ (0.228, 15.696) | 1.141^*^ (0.023, 57.571) | 1.00 |

**Table 7.7 Serious adverse events, approximate 36–39-week risk window**

| **OR（95% CI）** | **Placebo** | **Atacicept 150 mg** | **Cemdisiran 600 mg** | **Telitacicept 240 mg** |
| --- | --- | --- | --- | --- |
| **Placebo** | 1.00 |  |  |  |
| **Atacicept 150 mg** | 0.415^*^ (0.056, 3.081) | 1.00 |  |  |
| **Cemdisiran 600 mg** | 1.326^*^ (0.042, 41.440) | 3.191^*^ (0.059, 171.330) | 1.00 |  |
| **Telitacicept 240 mg** | 0.314^*^ (0.105, 0.938) | 0.756^*^ (0.077, 7.414) | 0.237^*^ (0.006, 8.775) | 1.00 |

Note: Cells show the effect of the row treatment relative to the column treatment and its 95% CI; all comparisons between active treatments are indirect comparisons connected through a common placebo. GMR, HR, and OR <1 and eGFR MD >0 favor the row treatment.

CINeMA certainty: **** high, *** moderate, ** low, * very low; asterisks do not indicate statistical significance. Proteinuria intervals are nominal 95% CIs; all seven active treatment–placebo comparisons in the primary window had P >0.05 after Holm adjustment. Kidney function and composite kidney outcomes were synthesized exploratorily using common-effect models; SAEs were analyzed using a Firth penalized-likelihood logistic model.

Dosing regimens: atacicept 150 mg subcutaneously once weekly; atrasentan 0.75 mg orally once daily; cemdisiran 600 mg subcutaneously every 4 weeks; iptacopan 200 mg orally twice daily; ravulizumab intravenously using weight-based dosing; SC0062 20 mg orally once daily; sibeprenlimab 400 mg subcutaneously or 4 mg/kg intravenously, both every 4 weeks; targeted-release budesonide 16 mg orally once daily; telitacicept 240 mg subcutaneously once weekly. All groups received background supportive treatment specified in the original trial.

Abbreviations: GMR, geometric mean ratio; MD, mean difference; HR, hazard ratio; OR, odds ratio; CI, confidence interval; eGFR, estimated glomerular filtration rate; SC, subcutaneous administration; IV, intravenous administration; TRF, targeted release. The unit for MD in total eGFR slope is mL/min/1.73 m²/year.

**Table 7.8 Holm adjustment for comparisons of active treatments with concurrent controls for proteinuria in the primary window**

| **Active treatment (vs trial-defined control)** | **Nominal P value** | **Holm-adjusted P value** |
| --- | --- | --- |
| Atacicept 150 mg | 0.0278 | 0.0834 |
| Atrasentan 0.75 mg | 0.0173 | 0.0782 |
| Cemdisiran 600 mg | 0.2449 | 0.2449 |
| Iptacopan 200 mg twice daily | 0.0156 | 0.0782 |
| Sibeprenlimab 400 mg subcutaneous | 0.0105 | 0.0647 |
| Targeted-release budesonide 16 mg | 0.0285 | 0.0834 |
| Telitacicept 240 mg | 0.0092 | 0.0647 |

Note: Two-sided t tests, residual df=2. All Holm-adjusted P values exceed 0.05.

**Table 7.9 Event components and confirmation criteria for composite kidney outcomes**

| **Study** | **eGFR decline** | **Persistence/confirmation criteria** | **Kidney failure component** | **Death component** |
| --- | --- | --- | --- | --- |
| Lafayette 2023 NefIgArd | ≥30% | Usually confirmed twice ≥4 weeks apart; exceptions include the final visit | Sustained eGFR<15; dialysis for ≥1 month; transplantation | Death due to kidney disease (included in kidney failure) |
| Heerspink 2026 ALIGN | ≥30% | ≥30 days | eGFR<15 sustained for ≥30 days; cumulative dialysis for ≥30 days; transplantation | All-cause death |
| Barratt 2026 APPLAUSE-IgAN | ≥30% | ≥4 weeks | eGFR<15 sustained for ≥4 weeks; dialysis for ≥4 weeks; transplantation | Death due to kidney disease (included in kidney failure) |

Note: NefIgArd generally required confirmation by two eGFR measurements at least 4 weeks apart; exceptions to confirmation applied in circumstances such as the final 24-month visit. The ALIGN composite outcome, which includes all-cause death, is presented separately.

#### **7.3 Network plots and forest plots**

**Figure 7.1 Evidence network for proteinuria in the primary window (32–40 weeks)**

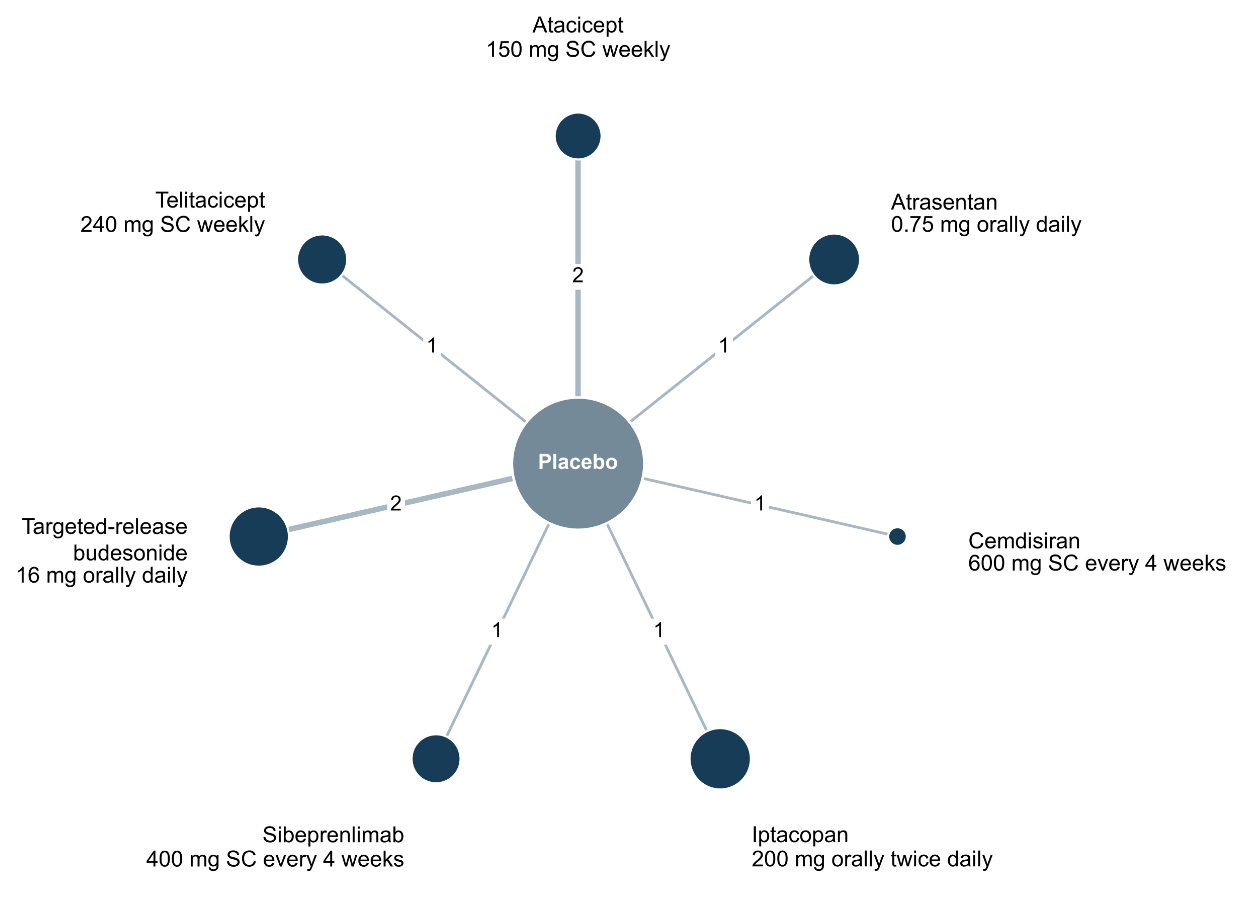

Note: The network includes 9 randomized controlled trials and 2,218 participants, with 8 nodes and 7 direct comparison edges. Node area is proportional to the number of participants analyzed in the corresponding treatment arms; line width and the numbers on the edges indicate the number of trials providing direct comparisons. All active regimens were compared with concurrent placebo; both groups received background supportive treatment as specified in the original trial. SC, subcutaneous.

**Figure 7.2 Evidence network for proteinuria in the broad window (24–52 weeks)**

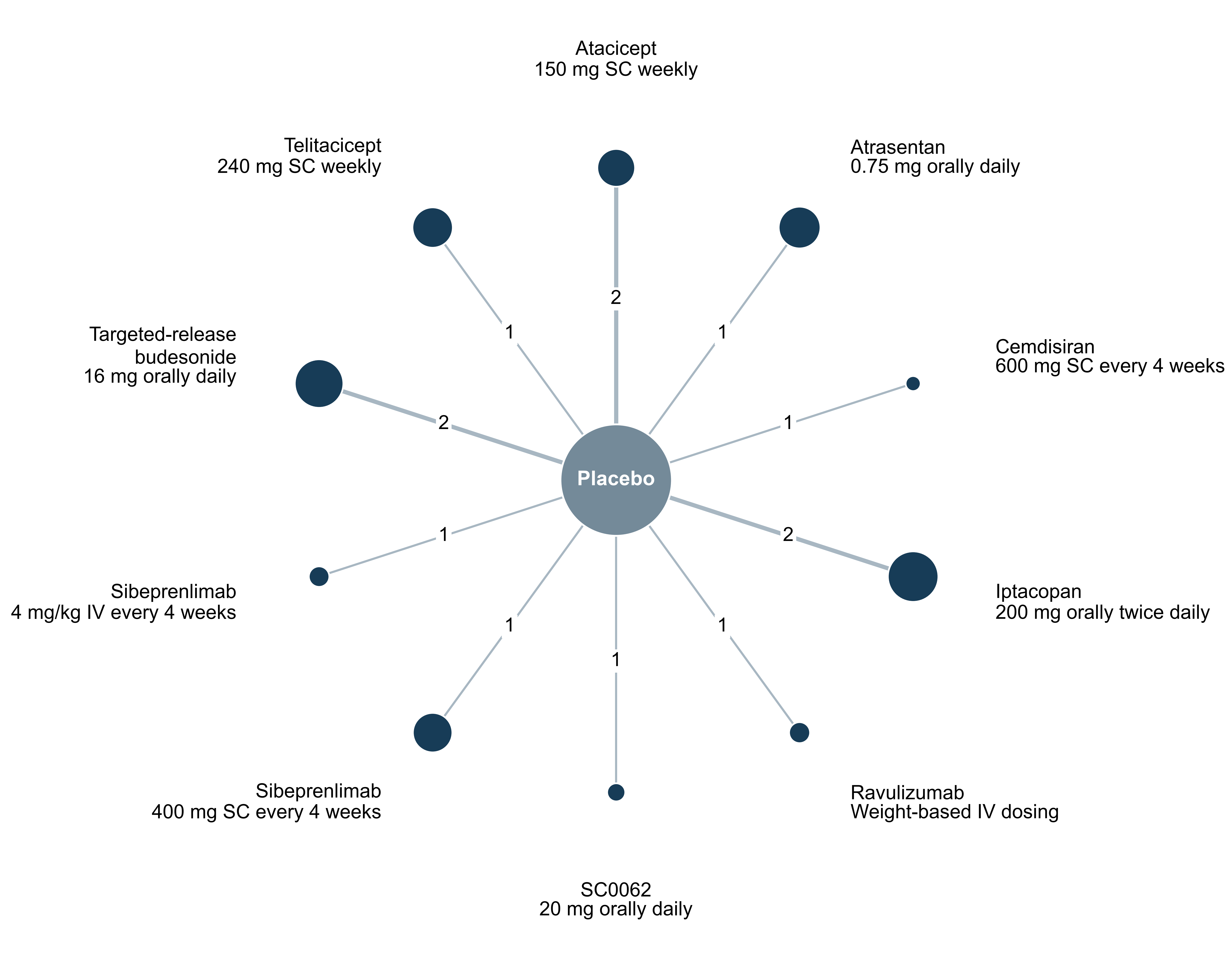

Note: A total of 13 randomized controlled trials involving 2,451 participants were included, comprising 11 nodes and 10 direct-comparison edges. Node area is proportional to the number of participants analyzed in the corresponding treatment arms; line width and the numbers on the edges indicate the number of trials providing direct comparisons. All active regimens were compared with concurrent placebo in the respective trials, and the network contained no closed loops; both groups received background supportive therapy as specified in the original trials. SC, subcutaneous administration; IV, intravenous administration.

**Figure 7.3. Forest plot of proteinuria in the primary window (32–40 weeks)**

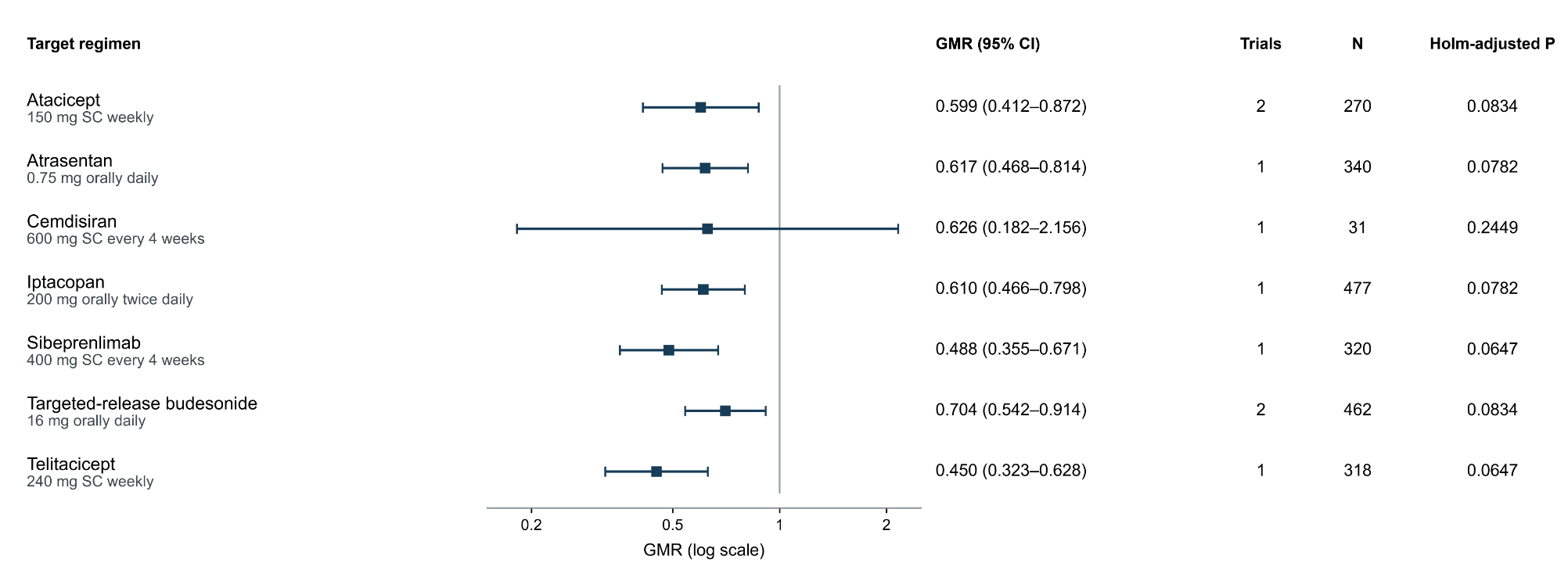

Note: A REML random-effects model with nominal t-distribution-based 95% confidence intervals was used (residual df = 2). GMR < 1 favors active treatment. The P values shown are Holm-adjusted P values for the 7 active treatment–placebo comparisons in the primary window. N is the sum of the numbers analyzed in the target treatment arm and the concurrent control arm. GMR, geometric mean ratio; CI, confidence interval; SC, subcutaneous administration.

**Figure 7.4. Forest plot of proteinuria in the broad window (24–52 weeks)**

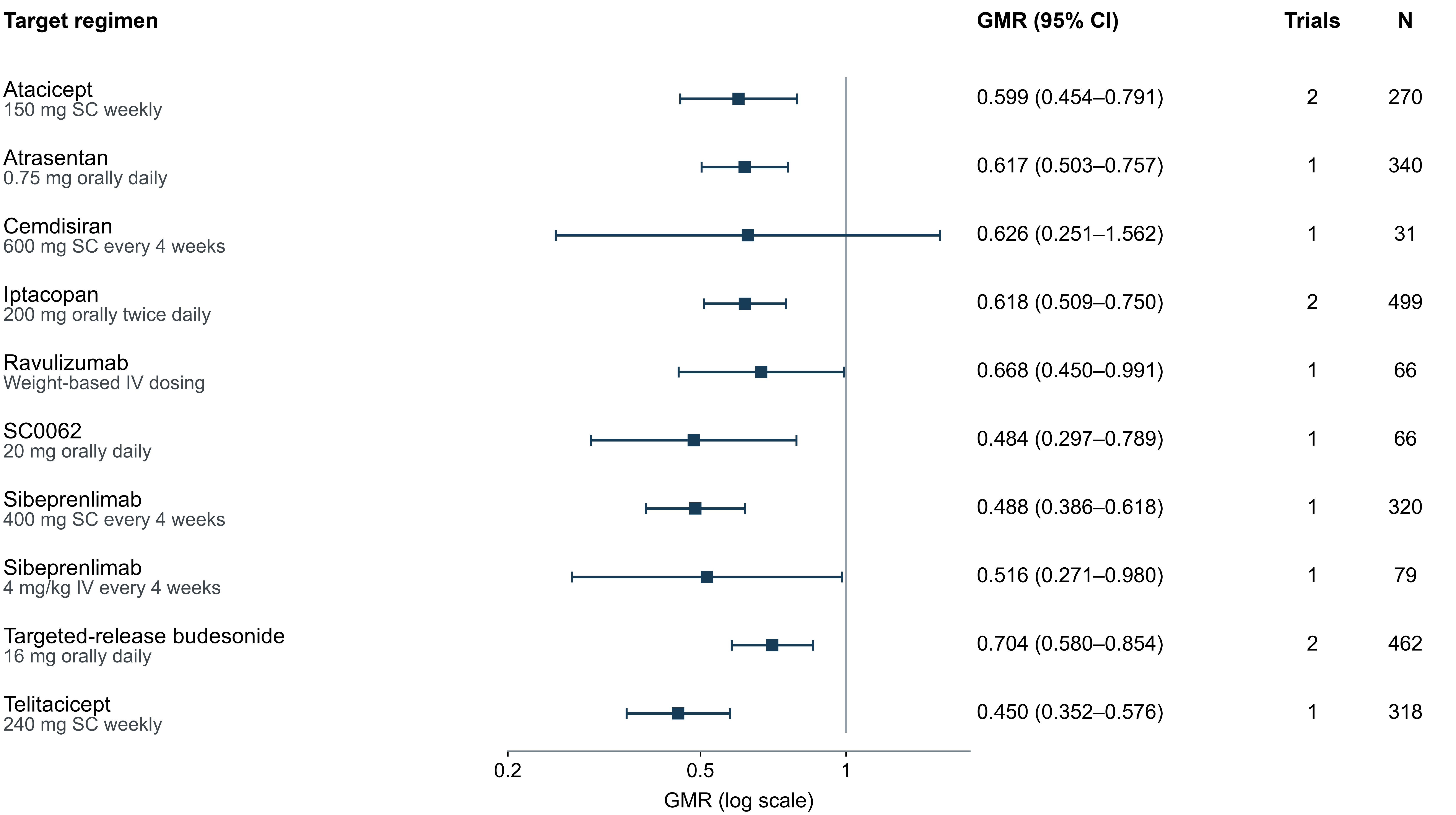

Note: GMRs and nominal 95% confidence intervals for each regimen versus concurrent placebo are shown. A total of 13 randomized controlled trials involving 2,451 participants were included; a REML random-effects model with t-distribution-based intervals was used (residual df = 3). GMR < 1 favors active treatment. N is the sum of the numbers analyzed in the target treatment arm and the concurrent control arm. GMR, geometric mean ratio; CI, confidence interval; SC, subcutaneous administration; IV, intravenous administration.

**Figure 7.5. Forest plot of annualized total eGFR slope**

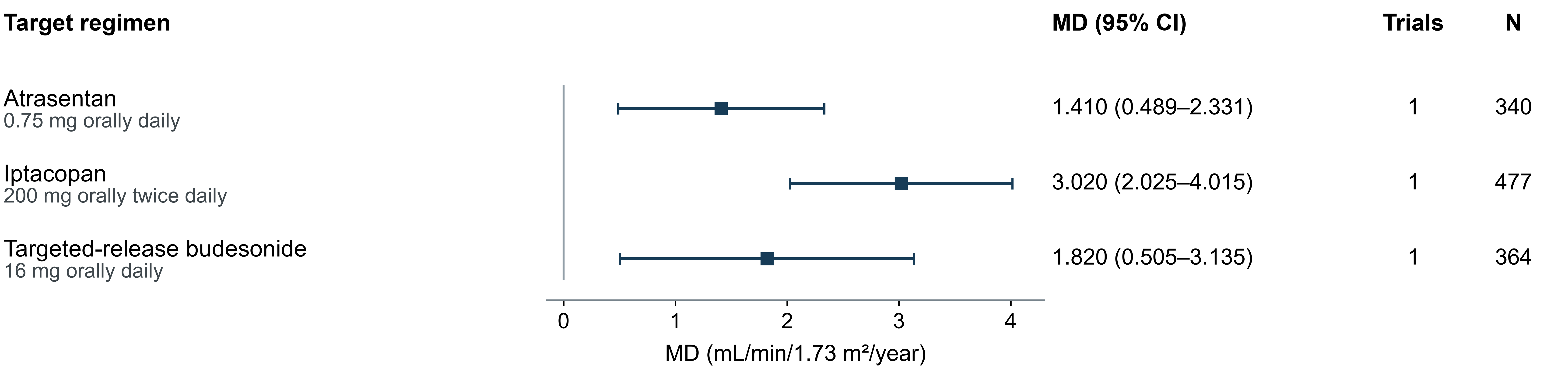

Note: Mean differences in annualized total eGFR slope and 95% confidence intervals for each regimen versus concurrent placebo are shown. A total of 3 randomized controlled trials involving 1,181 participants were included; a common-effect model with conditional normal intervals was used (residual df = 0). Slopes were calculated from randomization; atrasentan was evaluated through 136 weeks, and iptacopan and targeted-release budesonide through 104 weeks. MD > 0 indicates slower eGFR decline in the active treatment group. N is the sum of the numbers analyzed in the target treatment arm and the concurrent control arm. eGFR, estimated glomerular filtration rate; MD, mean difference; CI, confidence interval.

**Figure 7.6. Forest plot of composite kidney outcomes**

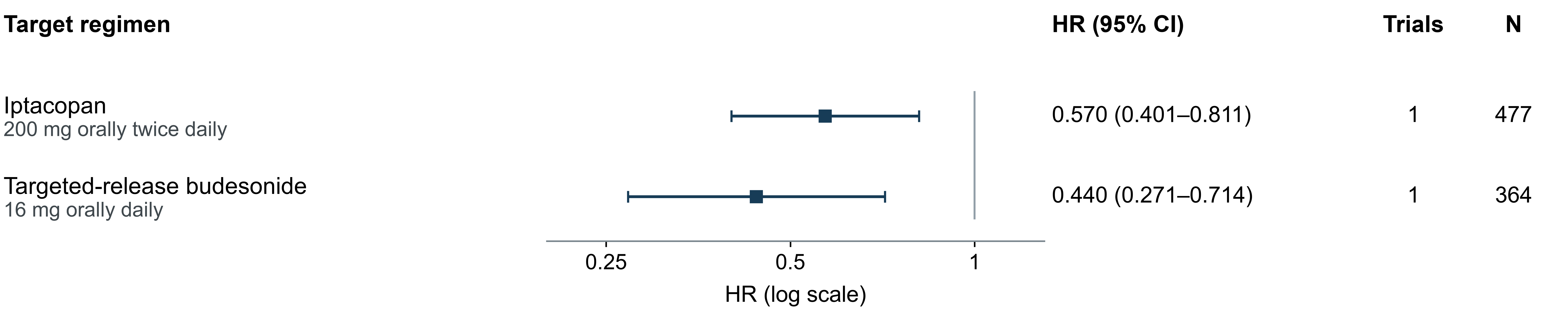

Note: HRs and 95% confidence intervals for each regimen versus concurrent placebo are shown. A total of 2 randomized controlled trials involving 841 participants were included, with assessment through 104 weeks; an exploratory common-effect model with conditional normal intervals was used (residual df = 0). HR < 1 favors active treatment; endpoint components, confirmation criteria, and handling of intercurrent events are presented in the corresponding tables. N is the sum of the numbers analyzed in the target treatment arm and the concurrent control arm. HR, hazard ratio; CI, confidence interval.

### **APPENDIX 8 Sensitivity and supplementary analyses**

#### **8.1 Proteinuria and other supplementary analyses**

**Table 8.1. Scope of sensitivity and supplementary analyses**

| **Analytical objective** | **Restriction or scenario** | **Scope** | **Limits of interpretation** |
| --- | --- | --- | --- |
| Time window | Primary window, 32–40 weeks; broad window, 24–52 weeks | Primary window, 9 RCTs; broad window, 13 RCTs | The broad window expands the assessment time points and treatment nodes |
| Heterogeneity | Common-effect reference; fixed-τ scenarios | Studies and nodes remain unchanged | Fixed τ represents a hypothetical scenario |
| Risk of bias | Results at low risk of bias according to RoB 2 only | Primary window, 3 RCTs; broad window, 4 RCTs | Common-effect subnetwork, residual df = 0 |
| Reporting stage | Exclusion of interim reports | VISIONARY, phase III ORIGIN, TELIGAN | A node is no longer estimable when its sole contributing study is removed |
| Time-window boundaries | Exclusion of the 32-/40-week boundaries | Cemdisiran、VISIONARY | Examines the impact of the primary-window boundary selection |
| Sparse events | LRP and MH without continuity correction | Separate analyses of the early and late windows | Estimability of zero-event comparisons varies by method |

**Table 8.2. Model sensitivity analyses of proteinuria effect estimates**

A Proteinuria primary window (32–40 weeks)

GMR (95% CI) for each regimen versus concurrent placebo

| **Treatment regimen** | **Primary analysis: random-effects REML , t distribution (df = 2)** | **Common effect,  τ = 0 , normal distribution** | **Fixed heterogeneity,  τ = 0.10 , normal distribution** | **Fixed heterogeneity,  τ = 0.20 , normal distribution** |
| --- | --- | --- | --- | --- |
| Atacicept 150 mg | 0.599 (0.412–0.872) | 0.599 (0.505–0.711) | 0.605 (0.482–0.758) | 0.610 (0.437–0.852) |
| Atrasentan 0.75 mg | 0.617 (0.468–0.814) | 0.617 (0.544–0.700) | 0.617 (0.489–0.779) | 0.617 (0.409–0.931) |
| Cemdisiran 600 mg | 0.626 (0.182–2.156) | 0.626 (0.357–1.100) | 0.626 (0.345–1.137) | 0.626 (0.315–1.244) |
| Iptacopan 200 mg  twice daily | 0.610 (0.466–0.798) | 0.610 (0.540–0.690) | 0.610 (0.484–0.769) | 0.610 (0.405–0.920) |
| Sibeprenlimab 400 mg  subcutaneously | 0.488 (0.355–0.671) | 0.488 (0.422–0.564) | 0.488 (0.382–0.623) | 0.488 (0.321–0.741) |
| Targeted-release budesonide 16 mg | 0.704 (0.542–0.914) | 0.704 (0.625–0.793) | 0.706 (0.583–0.855) | 0.707 (0.519–0.965) |
| Telitacicept 240 mg | 0.450 (0.323–0.628) | 0.450 (0.387–0.524) | 0.450 (0.351–0.577) | 0.450 (0.296–0.685) |

B Proteinuria broad window (24–52 weeks)

GMR (95% CI) for each regimen versus concurrent placebo

| **Treatment regimen** | **Primary analysis: random-effects REML , t distribution (df = 3)** | **Common effect,  τ = 0 , normal distribution** | **Fixed heterogeneity,  τ = 0.10 , normal distribution** | **Fixed heterogeneity,  τ = 0.20 , normal distribution** |
| --- | --- | --- | --- | --- |
| Atacicept 150 mg | 0.599 (0.454–0.791) | 0.599 (0.505–0.711) | 0.605 (0.482–0.758) | 0.610 (0.437–0.852) |
| Atrasentan 0.75 mg | 0.617 (0.503–0.757) | 0.617 (0.544–0.700) | 0.617 (0.489–0.779) | 0.617 (0.409–0.931) |
| Cemdisiran 600 mg | 0.626 (0.251–1.562) | 0.626 (0.357–1.100) | 0.626 (0.345–1.137) | 0.626 (0.315–1.244) |
| Iptacopan 200 mg  twice daily | 0.618 (0.509–0.750) | 0.618 (0.548–0.696) | 0.633 (0.510–0.785) | 0.656 (0.462–0.932) |
| Ravulizumab , weight-based dosing | 0.668 (0.450–0.991) | 0.668 (0.524–0.851) | 0.668 (0.489–0.913) | 0.668 (0.421–1.059) |
| SC0062 20 mg | 0.484 (0.297–0.789) | 0.484 (0.358–0.654) | 0.484 (0.338–0.693) | 0.484 (0.295–0.794) |
| Sibeprenlimab 400 mg  subcutaneously | 0.488 (0.386–0.618) | 0.488 (0.422–0.564) | 0.488 (0.382–0.623) | 0.488 (0.321–0.741) |
| Sibeprenlimab 4 mg/kg  intravenously | 0.516 (0.271–0.980) | 0.516 (0.347–0.766) | 0.516 (0.332–0.801) | 0.516 (0.295–0.900) |
| Targeted-release budesonide 16 mg | 0.704 (0.580–0.854) | 0.704 (0.625–0.793) | 0.706 (0.583–0.855) | 0.707 (0.519–0.965) |
| Telitacicept 240 mg | 0.450 (0.352–0.576) | 0.450 (0.387–0.524) | 0.450 (0.351–0.577) | 0.450 (0.296–0.685) |

Note: GMR < 1 favors active treatment; all intervals are nominal 95% confidence intervals. The primary analysis used REML to estimate the common between-study variance τ² and constructed intervals based on the t distribution; the common-effect and fixed-τ scenarios used conditional normal intervals. τ is the between-study standard deviation on the log GMR scale. τ = 0.10 and 0.20 correspond to τ² = 0.01 and 0.04, respectively, and are specified scenario values for the sensitivity analyses. Differences in intervals between the primary analysis and the other scenarios reflect the effects of both the heterogeneity specification and the interval calculation method; intervals in the fixed-τ scenarios are conditional on the specified τ. GMR, geometric mean ratio; CI, confidence interval; REML, restricted maximum likelihood; df, degrees of freedom.

**Table 8.3. Proteinuria analyses restricted to results at low risk of bias**

| **Analysis set** | **Active treatment** | **GMR（95% CI）** |
| --- | --- | --- |
| **Primary window: 3 RCTs, 841 participants; residual df = 0** | | |
| Primary window, 32–40 weeks | Atacicept 150 mg | 0.582 (0.477–0.711) |
| Primary window, 32–40 weeks | Sibeprenlimab 400 mg subcutaneous | 0.488 (0.422–0.564) |
| Primary window, 32–40 weeks | Telitacicept 240 mg | 0.450 (0.387–0.524) |
| **Broad window: 4 RCTs, 907 participants; residual df = 0** | | |
| Broad window, 24–52 weeks | Atacicept 150 mg | 0.582 (0.477–0.711) |
| Broad window, 24–52 weeks | Ravulizumab, weight-based dosing | 0.668 (0.524–0.851) |
| Broad window, 24–52 weeks | Sibeprenlimab 400 mg subcutaneous | 0.488 (0.422–0.564) |
| Broad window, 24–52 weeks | Telitacicept 240 mg | 0.450 (0.387–0.524) |

Note: Only results at low risk of bias according to RoB 2 for the corresponding outcome were included; a common-effect model with conditional normal intervals was used

**Table 8.4. Analyses excluding interim and stage-specific reports and time-window boundaries**

| **Active treatment** | **Original model  GMR (95% CI)** | **Excluding interim reportsᵃ  GMR (95% CI)** | **Further excluding Part Aᵇ  GMR (95% CI)** | **Excluding 32-/40-week boundariesᶜ  GMR (95% CI)** |
| --- | --- | --- | --- | --- |
| **Primary window, 32–40 weeks** | | | | |
| Atacicept 150 mg | 0.599 (0.412–0.872) | 0.650 (0.076–5.546) | 0.650 (0.076–5.546) | 0.599 (0.412–0.872) |
| Atrasentan 0.75 mg | 0.617 (0.468–0.814) | 0.617 (0.272–1.397) | 0.617 (0.272–1.397) | 0.617 (0.468–0.814) |
| Cemdisiran 600 mg | 0.626 (0.182–2.156) | 0.626 (0.016–24.092) | 0.626 (0.016–24.092) | Not estimable |
| Iptacopan 200 mg twice daily | 0.610 (0.466–0.798) | 0.610 (0.276–1.350) | 0.610 (0.276–1.350) | 0.610 (0.466–0.798) |
| Sibeprenlimab 400 mg subcutaneous | 0.488 (0.355–0.671) | Not estimable | Not estimable | Not estimable |
| Targeted-release budesonide 16 mg | 0.704 (0.542–0.914) | 0.704 (0.326–1.521) | 0.704 (0.326–1.521) | 0.704 (0.542–0.914) |
| Telitacicept 240 mg | 0.450 (0.323–0.628) | 0.450 (0.168–1.202) | Not estimable | 0.450 (0.323–0.628) |
| **Broad window, 24–52 weeks** | | | | |
| Atacicept 150 mg | 0.599 (0.454–0.791) | 0.650 (0.314–1.343) | 0.650 (0.314–1.343) | 0.599 (0.454–0.791) |
| Atrasentan 0.75 mg | 0.617 (0.503–0.757) | 0.617 (0.468–0.814) | 0.617 (0.468–0.814) | 0.617 (0.503–0.757) |
| Cemdisiran 600 mg | 0.626 (0.251–1.562) | 0.626 (0.182–2.156) | 0.626 (0.182–2.156) | Not estimable |
| Iptacopan 200 mg twice daily | 0.618 (0.509–0.750) | 0.618 (0.475–0.803) | 0.618 (0.475–0.803) | 0.618 (0.509–0.750) |
| Ravulizumab, weight-based dosing | 0.668 (0.450–0.991) | 0.668 (0.392–1.138) | 0.668 (0.392–1.138) | 0.668 (0.450–0.991) |
| SC0062 20 mg | 0.484 (0.297–0.789) | 0.484 (0.250–0.938) | 0.484 (0.250–0.938) | 0.484 (0.297–0.789) |
| Sibeprenlimab 400 mg subcutaneous | 0.488 (0.386–0.618) | Not estimable | Not estimable | Not estimable |
| Sibeprenlimab 4 mg/kg intravenous | 0.516 (0.271–0.980) | 0.516 (0.216–1.228) | 0.516 (0.216–1.228) | 0.516 (0.271–0.980) |
| Targeted-release budesonide 16 mg | 0.704 (0.580–0.854) | 0.704 (0.542–0.914) | 0.704 (0.542–0.914) | 0.704 (0.580–0.854) |
| Telitacicept 240 mg | 0.450 (0.352–0.576) | 0.450 (0.323–0.628) | Not estimable | 0.450 (0.352–0.576) |

Note: ᵃ Interim reports from Perkovic 2026 VISIONARY and Lafayette 2026 phase III ORIGIN were excluded. ᵇ The Lv 2026 TELIGAN Part A report was further excluded. ᶜ Barratt 2024 Cemdisiran (32 weeks) and VISIONARY (40 weeks) were excluded. After excluding interim reports/further excluding Part A/excluding boundaries, the primary window retained 7/6/7 RCTs, with residual df of 1/1/2, respectively; the broad window retained 11/10/11 RCTs, with residual df of 2/2/3, respectively.

#### **8.2 Model sensitivity analyses of serious adverse events**

**Table 8.5. LRP and Mantel–Haenszel estimates for serious adverse events**

| **Treatment and risk window** | **Participants with events/participants analyzed in the active arm** | **Participants with events/participants analyzed in the control arm** | **LRP primary model  OR (95% CI)** | **MH without continuity correction  OR (95% CI)** |
| --- | --- | --- | --- | --- |
| **Serious adverse events, 24–26 weeks** | | | | |
| Ravulizumab, weight-based dosing | 1/43 | 0/23 | 1.659 (0.061–45.047) | No finite estimate¹ |
| SC0062 20 mg | 2/31 | 1/34 | 1.893 (0.228–15.696) | 2.276 (0.196–26.420) |
| **Serious adverse events, 36–39 weeks** | | | | |
| Atacicept 150 mg | 1/33 | 3/34 | 0.415 (0.056–3.081) | 0.323 (0.032–3.274) |
| Cemdisiran 600 mg | 1/22 | 0/9 | 1.326 (0.042–41.440) | No finite estimate¹ |
| Telitacicept 240 mg | 4/159 | 13/159 | 0.314 (0.105–0.938) | 0.290 (0.092–0.909) |

*LRP, penalized-likelihood logistic regression; MH, Mantel–Haenszel common-effect method; neither method uses a 0.5 continuity correction. ¹ Ravulizumab and Cemdisiran are single-study edges with zero events in the control arm*

### **APPENDIX 9 Raw serious adverse event data and risk windows**

**Table 9.1. Raw serious adverse event counts and collection periods**

| **Study** | **Target treatment and concurrent control** | **Safety analysis population** | **Actual collection period and cutoff** | **Target arm, ≥1 SAE, n/N**  **（%）** | **Control arm, ≥1 SAE, n/N**  **（%）** |
| --- | --- | --- | --- | --- | --- |
| *Heerspink 2025 SC0062* | SC0062 20 mg orally once daily; concurrent matching placebo | Received ≥1 dose: 31/34 | 24 weeks of treatment and 4 weeks of safety follow-up after the last dose | 2/31（6.5%） | 1/34（2.9%） |
| *Lafayette 2025 SANCTUARY* | Ravulizumab, weight-based intravenous loading and maintenance doses; concurrent matching placebo | Randomized and received ≥1 dose: 43/23 | Initial 26-week controlled period | 1/43（2.3%） | 0/23（0.0%） |
| *Barratt 2024 Cemdisiran* | Cemdisiran 600 mg subcutaneously every 4 weeks; concurrent matching placebo | Double-blind-period safety analysis set: 22/9 | 36-week double-blind period; first dose to 28 days after the last dose | 1/22（4.5%） | 0/9（0.0%） |
| *Lafayette 2024  ORIGIN phase IIb* | Atacicept 150 mg subcutaneously once weekly; concurrent matching placebo | Safety analysis set: 33/34 | Through 36 weeks | 1/33（3.0%） | 3/34（8.8%） |
| *Lv 2026 TELIGAN* | Telitacicept 240 mg subcutaneously once weekly; concurrent matching placebo | Received ≥1 dose and had safety data: 159/159 | First dose to before dosing at week 40 (nominally 39 weeks) | 4/159（2.5%） | 13/159（8.2%） |
| *Fellström 2017 NEFIGAN* | TRF-budesonide 16 mg orally once daily; concurrent matching placebo | Received ≥1 dose: 49/50 | 9 months of treatment + 3 months of follow-up | Overall number of participants not obtained | Overall number of participants not obtained |
| *Lafayette 2023 NefIgArd* | Nefecon 16 mg orally once daily; concurrent matching placebo | Full analysis set in the original report: 182/182; actually received ≥1 dose: 180/179 | First dose to 14 days after the last dose (including tapering) | 18/182（9.9%） | 9/182（4.9%） |
| *Heerspink 2026 ALIGN* | Atrasentan 0.75 mg orally once daily; concurrent matching placebo | Randomized and received ≥1 dose: 169/170 | First dose to 30 days after the last dose (132 weeks of treatment) | 24/169（14.2%） | 26/170（15.3%） |
| *Barratt 2026 APPLAUSE-IgAN* | Iptacopan 200 mg orally twice daily; concurrent matching placebo | Received ≥1 dose: 238/239 | Cumulative over 24 months | 29/238（12.2%） | 28/239（11.7%） |
| *Lv 2023  Telitacicept phase II* | Telitacicept 240 mg subcutaneously once weekly; concurrent matching placebo | Safety reporting population: 14/14 | Informed consent to 28 days after the last dose (24 weeks of treatment) | 2/14（14.3%） | 1/14（7.1%） |
| *Mathur 2024 ENVISION* | Sibeprenlimab 4 mg/kg intravenously every 4 weeks; concurrent matching placebo | Randomized and received study drug: 41/38 | 16 months, including post-treatment follow-up | 2/41（4.9%） | 2/38（5.3%） |
| *Perkovic 2026 VISIONARY* | Sibeprenlimab 400 mg subcutaneously every 4 weeks; concurrent matching placebo | Received ≥1 dose: 259/251 | Cumulative at interim analysis, with unequal follow-up | 9/259（3.5%） | 11/251（4.4%） |
| *Lafayette 2026  ORIGIN phase III* | Atacicept 150 mg subcutaneously once weekly; concurrent matching placebo | Randomized and received ≥1 dose: 214/214 | Cumulative at interim analysis, with unequal exposure | 1/214（0.5%） | 11/214（5.1%） |
| *Floege 2025 IGNAZ* | Felzartamab, 9 weight-based intravenous doses; concurrent matching placebo | Received ≥1 dose, grouped by actual treatment: 13/12 | On-treatment period: first dose to 28 days after the last dose | 0/13（0.0%） | 0/12（0.0%） |
| *ARTEMIS-IgAN* | Narsoplimab 370 mg intravenous regimen; concurrent matching placebo | Registered SAE analysis population: 181/179 | Blinded period: first dose to a maximum of 96 weeks or the last visit | 22/181（12.2%） | 20/179（11.2%） |
| *ALXN2050 IgAN* | Vemircopan 180 mg orally twice daily; concurrent matching placebo | Participants with IgAN who received ≥1 dose: 26/26 | Days 1–813; entire period for the target arm, initial period for the control arm | 2/26（7.7%） | 2/26（7.7%） |
| *Zhang 2024 Iptacopan* | Iptacopan 200 mg orally twice daily; concurrent matching placebo | Separate safety population count for Part 2 unavailable | Treatment period to 7 days after the last dose (up to 197 days in Part 2) | Separate count/denominator not obtained | Separate count/denominator not obtained |

Note: SAE, serious adverse event; TEAE, adverse event that emerged or worsened during treatment. n/N represents the number of participants with at least one SAE and the corresponding analysis denominator in the original report; percentages were calculated from this fraction and rounded to 1 decimal place.

#### **9.1 Safety model plots**

**Figure 9.1. Primary and sensitivity analyses of serious adverse events at 24–26 weeks**

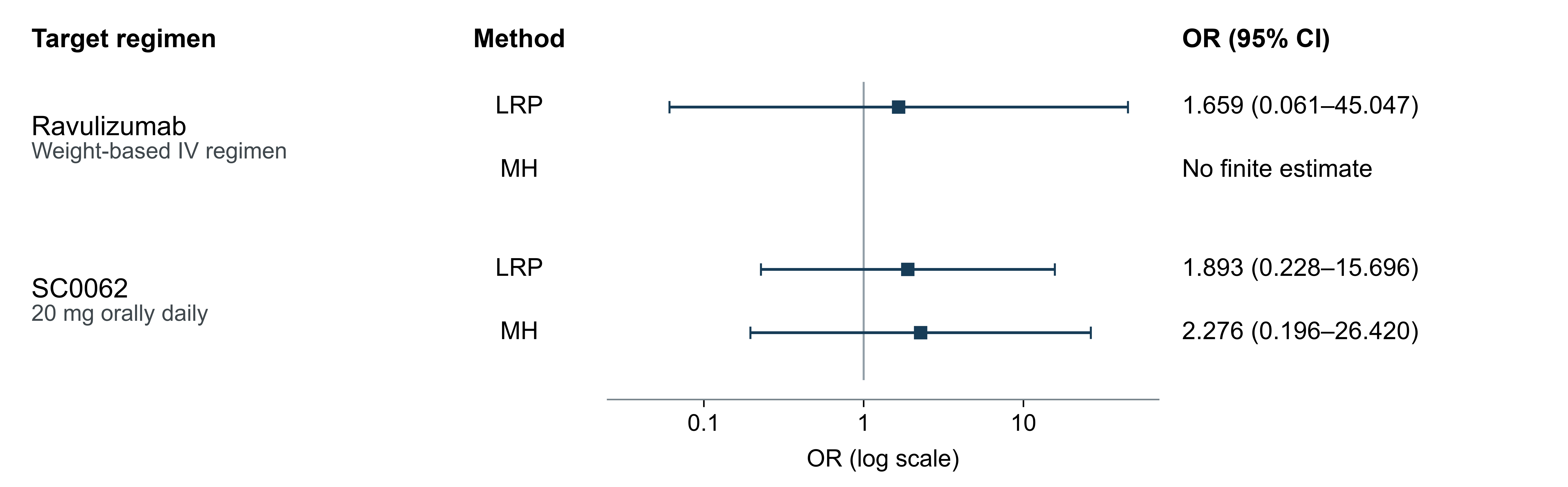

Note: Each regimen was compared with concurrent placebo. Squares and horizontal lines represent ORs and 95% CIs, respectively; the horizontal axis is logarithmic, and OR < 1 indicates lower odds of at least one serious adverse event in the active treatment group. LRP is the penalized-likelihood logistic regression common-effect primary model, with normal Wald intervals; MH is the Mantel–Haenszel sensitivity analysis without continuity correction. The ravulizumab control group had zero events, and MH did not yield a finite estimate. OR, odds ratio; CI, confidence interval; IV, intravenous administration.

**Figure 9.2. Primary and sensitivity analyses of serious adverse events at 36–39 weeks**

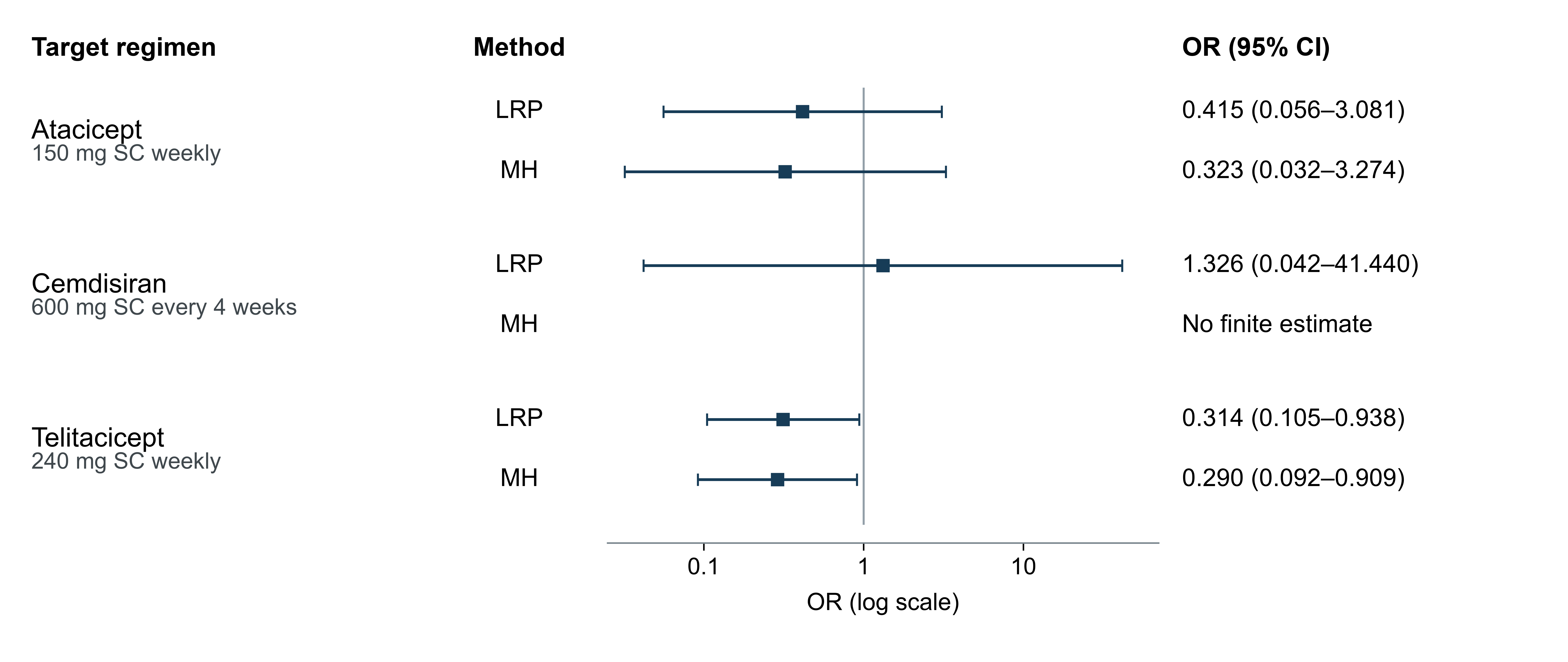

Note: Each regimen was compared with concurrent placebo. Squares and horizontal lines represent ORs and 95% CIs, respectively; the horizontal axis is logarithmic, and OR < 1 indicates lower odds of at least one serious adverse event in the active treatment group. LRP is the penalized-likelihood logistic regression common-effect primary model, with normal Wald intervals; MH is the Mantel–Haenszel sensitivity analysis without continuity correction. The cemdisiran control group had zero events, and MH did not yield a finite estimate. OR, odds ratio; CI, confidence interval; SC, subcutaneous administration.

### **APPENDIX 10 Subsequent trials and long-term follow-up**

The following table lists new randomized trials relevant to this review, pending subsequent kidney outcomes from included trials, and candidate trials at the boundaries of the review scope or with eligibility yet to be clarified.

**Table 10.1. Registration information and pending outcomes of relevant randomized trials**

| **Registration number and trial name** | **Region and study population** | **Intervention and comparator** | **Registered enrollment** | **Primary and key kidney outcomes and assessment time points** | **Registered status and primary completion date** |
| --- | --- | --- | --- | --- | --- |
| **A New relevant randomized trials** | | | | | |
| BEYOND NCT05852938 | Multinational phase III; ≥18 years; biopsy-confirmed primary IgAN; persistent proteinuria | Zigakibart 600 mg subcutaneously every 2 weeks for 104 weeks; matching placebo | 383 (actual) | Change in eGFR: 104 weeks (primary); UPCR: 40 weeks; eGFR slope and composite kidney events: 104 weeks | Active, not recruiting; primary completion: 2027-08-10 (estimated) |
| IMAGINATION NCT05797610 | Multinational phase III; ≥18 years; biopsy-confirmed primary IgAN; eGFR ≥20; UPCR ≥1 g/g or urinary protein ≥1 g/d and UPCR ≥0.8 | Sefaxersen subcutaneously, with initial doses on days 1, 15, and 29, then every 4 weeks through week 105; matching placebo | 459 (actual) | 24-h UPCR: 37 weeks (primary); eGFR slope: 105 weeks; composite kidney failure events: up to approximately 36 months | Active, not recruiting; primary completion: 2028-05-25 (estimated) |
| RAINIER NCT06564142 | Multinational phase III; adults; biopsy-confirmed IgAN; eGFR ≥30; urinary protein ≥1 g/d or 24-h UPCR ≥0.75 g/g | Povetacicept subcutaneously; matching placebo | 605 (actual) | 24-h UPCR: 36 weeks and total eGFR slope: 104 weeks (both primary); change in eGFR and kidney disease progression events: 104 weeks | Active, not recruiting; primary completion: 2028-01-30 (estimated) |
| PREVAIL NCT06935357 | Multinational phase III; adults; biopsy-confirmed IgAN; main cohort eGFR ≥30; additional cohort with eGFR 20–<30 | Felzartamab intravenously, with 24 weeks of treatment and follow-up through 104 weeks; matching placebo | 454 (estimated) | Percentage change in UPCR: 36 weeks (primary); change in eGFR: 52 and 104 weeks; composite kidney events and safety: 104 weeks | Recruiting; primary completion: 2027-05-31 (estimated) |
| I CAN NCT06291376 | Multinational phase III; adults; biopsy-confirmed IgAN; main cohort eGFR ≥30; additional advanced kidney disease cohort with eGFR 20–29 | Ravulizumab, weight-based intravenous loading dose, maintenance dosing from day 15 and every 8 weeks thereafter; matching placebo | 584 (actual) | 24-h UPCR: 34 weeks (interim primary); change in eGFR: 106 weeks (final primary); total eGFR slope: 50 weeks; composite kidney events: 106 weeks | Active, not recruiting; primary completion: 2026-01-05 (actual); study completion: 2030-07-18 (estimated) |
| SUCCESS-1 NCT06819826 | Phase III in China; ≥18 years; biopsy-confirmed IgAN; 30 ≤eGFR <90; UPCR ≥0.75 g/g or urinary protein ≥1 g/d; stable background therapy for ≥12 weeks | SC0062 20 mg orally once daily for 108 weeks; matching placebo | 360 (estimated) | 24-h UPCR: 36 weeks (primary); change in eGFR and total eGFR slope: 108 weeks | Recruiting; primary completion: 2026-12-31 (estimated) |
| **B Subsequent kidney outcomes from included trials** | | | | | |
| VISIONARY NCT05248646 | Multinational phase III; ≥18 years; primary IgAN; main cohort eGFR ≥30; additional exploratory cohort with low eGFR | Sibeprenlimab 400 mg subcutaneously every 4 weeks; matching placebo | 530 (actual) | 24-h UPCR at 9 months reported; total eGFR slope, change in eGFR, and composite kidney failure events at 24 months pending | Active, not recruiting; primary completion: 2026-05-13 (actual); study completion: 2027-01-19 (estimated) |
| ORIGIN 3 NCT04716231 | Multinational phase III; ≥18 years; biopsy-confirmed IgAN; eGFR ≥30; UPCR ≥1 g/g or urinary protein ≥1 g/d | Atacicept 150 mg subcutaneously once weekly; matching placebo; 104-week double-blind period followed by an open-label period | 428 (actual) | UPCR at 36 weeks reported; long-term kidney function results from the 104-week double-blind period pending; the subsequent 52-week open-label period and 26-week safety follow-up reported separately | Active, not recruiting; primary completion: 2025-05-15 (actual); study completion: 2028-07 (estimated) |
| TELIGAN NCT05799287 | Phase III in China; ≥18 years; primary IgAN; eGFR ≥30; persistent proteinuria | Telitacicept 240 mg subcutaneously; Part A, 39 once-weekly doses; Part B, 32 doses every 2 weeks; matching placebo | 318 (actual) | UPCR at 39 weeks reported; annualized eGFR slope at 104 weeks (Part B primary), sustained eGFR decline, and composite kidney events pending | Active, not recruiting; primary and study completion: 2026-06 (estimated) |
| **C Candidate trials at the boundaries of the review scope or with eligibility yet to be clarified** | | | | | |
| Mezagitamab TAK-079-3001 NCT06963827 | Multinational phase III; ≥18 years; biopsy-confirmed primary IgAN; main randomized cohort eGFR >30, UPCR ≥0.8 g/g or urinary protein ≥1 g/d; additional open-label cohort | Mezagitamab subcutaneously for approximately 22 weeks; matching placebo; 2:1 randomization, with follow-up through 104 weeks; additional open-label cohort | 347 (estimated) | 24-h UPCR: 36 weeks (primary); change in eGFR: 52 and 104 weeks; total eGFR slope, sustained eGFR decline, and composite kidney failure events: 104 weeks | Recruiting; primary completion: 2028-09-07 (estimated) |
| CM313-105101 NCT06830395 | Phase II in China; 18–75 years; biopsy-confirmed primary IgAN; eGFR ≥30; UPCR ≥0.75 g/g or urinary protein ≥1 g/d | CM313 subcutaneously at different doses and frequencies; the registry specifies matching placebo in Part B, with Part A recorded separately | 106 (estimated) | Proportion with adverse events: up to 108 weeks (registered primary outcome); the registry page does not list specific assessment time points for proteinuria/eGFR | Not yet recruiting; primary completion: 2027-03 (estimated) |

Note: Registration information is current as of October 8, 2026. The listed statuses and dates represent a snapshot of registry information. UPCR, urine protein-to-creatinine ratio; eGFR, estimated glomerular filtration rate, in mL/min/1.73 m².

#### **Methodological references**

6. Cochrane Handbook for Systematic Reviews of Interventions. Chapters 4, 6, 10, 11 and technical supplement to searching. https://training.cochrane.org/handbook

7. Chiocchia V, Holloway A, Salanti G. Semi-automated assessment of the risk of bias due to missing evidence in network meta-analysis: a guidance paper for the ROB-MEN web-application. BMC Med Res Methodol. (2023) 23:223. doi: 10.1186/s12874-023-02038-9.

8. ICH E9(R1). Addendum on estimands and sensitivity analysis in clinical trials. 2019.
